# Proteomics analysis of peripheral blood monocytes from patients in early dengue infection reveals potential markers of subsequent fluid leakage

**DOI:** 10.1101/2024.03.21.24304389

**Authors:** Nilanka Perera, Abhinav Kumar, Bevin Gangadharan, Diyanath Ranasinghe, Ananda Wijewickrama, Gathsaurie Neelika Malavige, Joanna L. Miller, Nicole Zitzmann

## Abstract

Infections caused by dengue virus (DENV) cause significant morbidity and mortality worldwide. The majority of patients have a mild course of dengue fever (DF) disease, however a proportion of infected individuals develop much more severe dengue haemorrhagic fever (DHF) resulting in circulatory collapse and multiorgan failure due to increased vascular permeability. Early detection of individuals likely to develop severe disease could lead to improved outcomes for patients, and help use healthcare resources more efficiently. At present there are no reliable markers during the earlier stages of infection that indicate which patients will go on to develop DHF. Our study was aimed at identifying proteins that are differentially regulated early during disease in peripheral blood monocytes (PBMC) of patients who subsequently develop DHF. Such proteins may also point at cellular pathways implicated in developing vascular leakage. PBMC were isolated from patients with a confirmed dengue infection, lysed and subjected to tandem mass tag mass spectrometry. One hundred and sixty proteins were differentially expressed in DENV-infected samples compared to healthy controls. These were mainly involved in type I interferon signaling, cytokine response, phagocytosis, haemostasis and cell adhesion. PBMC from DHF patients differentially expressed 90 proteins compared to individuals with DF; these were involved in down-regulation of platelet activation and aggregation, cell adhesion and cytoskeleton arrangement pathways. Proteins involved in oxidative stress and p38 MAPK signaling were upregulated in DHF samples during early infection compared to DF samples. The proteins reported here that are differentially regulated in PBMC early during infection could potentially serve as biomarkers to identify patients at risk of developing DHF at an early disease stage. This study also provides important observations on pathways implicated in severe DENV infection.

## Introduction

Dengue virus (DENV) infections are prevalent throughout the world with an estimated 390 million cases occurring every year (1). Asia harbors approximately seventy percent of this viral infection which causes significant morbidity and mortality (1). DENV infections account for 1.1 million disability-adjusted life-years globally (2). In the majority of cases dengue fever (DF) is characterized by an initial febrile phase with viraemia, followed by resolution of fever leading to recovery (3). A minority of DENV-infected individuals develop plasma leakage at the time of defervescence, known as the “critical phase” which typically lasts 24-48 hours. The increased vascular permeability causing plasma leakage is reflected by the development of serosal effusions and a rise in haematocrit (4). Such dengue haemorrhagic fever (DHF) patients, unless resuscitated with adequate fluids, develop shock leading to multiorgan failure and coagulopathy, which causes significant mortality. Patients with severe plasma leakage (leading to shock or respiratory distress), severe bleeding and severe organ impairment are considered as severe dengue patients (5). Prevalence of DHF varies from 6.3% in DENV-infected individuals (irrespective of symptoms) to 45.7% in symptomatic and hospitalized cases (6). There are no clinical or biochemical markers at present that accurately predict the development of DHF during early infection. Thus, stringent monitoring and regular bedside ultrasonography in healthcare facilities are required to identify patients developing DHF. Inability to triage patients during early infection results in overcrowding of health care facilities and increased utilization of resources.

The pathogenesis of plasma leakage is presumed to be due to an alteration in the endothelial glycocalyx. Studies suggest a possible role of DENV non-structural protein 1 (NS1) and its interactions with the endothelium (7,8), although exactly how this happens is unknown. The roles of many cytokines and vascular adhesion markers have been studied in severe disease, though with limited utility in clinical practice. In addition, increased endothelial cell permeability could be due to complex interactions between DENV-infected cells such as monocytes, platelets and the vascular endothelium. Some studies have explored the transcriptome of DENV-infected peripheral blood monocytes (PBMC) in an effort to identify differential gene signatures leading to severe disease (9–11). Although such analysis may provide useful insights, disease usually manifests on the protein rather than the gene level, and identification of key proteins is also more suitable for developing tests for relevant markers. To date, there are no data on the proteome of PBMCs of DENV-infected patients early during infection. The proteome of infected monocytes in patients with DHF during early infection could also provide valuable information on the pathways triggered at that point that may subsequently lead to plasma leakage. Here we report the results of a tandem mass tag (TMT) mass spectrometry experiment and comprehensive analysis aimed at identifying differentially regulated proteins in patients with DF compared to those who subsequently went on to develop DHF, using samples of healthy individuals as controls. The present study identifies previously under-recognized biological pathways and individual proteins potentially involved in causing plasma leakage, laying the foundation for future validation studies.

## Results

We recruited 25 patients (17 DF and 8 patients who subsequently developed DHF) to the study who fulfilled the selection criteria, and early during the course of the disease collected their PBMC to be lysed. The protein content of the lysates was quantified and the four DF and four DHF samples with the highest protein content were selected for subsequent TMT mass spectrometry experiments. PBMC lysates from two healthy controls, four DF and four DHF patients were analysed by TMT mass spectrometry. The sera of the selected patients were positive for DENV viral genome identified by qRT-PCR. All patients (n=8) were infected by DENV serotype 2. Patients who developed DHF had evidence of fluid leakage (pleural effusion and/or ascites) during their later course of illness. The characteristics of the patients are given in Table 1. DHF patients had a significantly lower platelet count over the course of illness. Other parameters were not significantly different between the two groups.

**Table 1:** Characteristics of the patients selected for the TMT mass spectrometry study.

| Clinical parameter | DF<br>n=4 | DHF<br>n=4 |
| --- | --- | --- |
| Age in years, mean (SD) | 25.7 (7.9) | 38.8 (8.8) |
| Males (n, %) | 2 (50) | 2 (50) |
| Laboratory investigations on admission in the febrile phase, mean (SD) |  |  |
| White cells ($\times 10^9/L$) | 6.2 (3.3) | 5.5 (1.46) |
| Platelets ($\times 10^9/L$) | 189 (82) | 193 (62) |
| Packed cell volume (%) | 39.9 (3.2) | 39.2 (5.1) |
| Highest/lowest values of laboratory investigations during the course of illness, mean (SD) |  |  |
| Lowest white cells ($\times 10^9/L$) | 2.7 (0.4) | 4.4 (1.5) |
| Lowest platelets ($\times 10^9/L$)* | 70.5 (37.2-157) | 13.5 (12.2-26) |
| Highest packed cell volume (%) | 42.2 (5.2) | 45.2 (7.5) |
| Liver enzymes, median (IQR) |  |  |
| AST (U/L) | 68.7 (30.4-157) | 65 (51.9-394.2) |
| ALT (U/L) | 65.1 (40.6-154.7) | 54.8 (28.9-339.5) |
| DENV diagnosis |  |  |
| Positive NS1 rapid kit | 4 (100) | 4 (100) |
| Virus detected by qRT-PCR | 4 (100) | 4 (100) |
| Viral load in the acute phase ($\log_{10}$ GE)* | 2.1 (1.5-5.0) | 1.9 (1.4-2.3) |

\*Values given as median (IQR)

The initial check for successful TMT labelling of the unfractionated PBMC lysate digest revealed 1018 proteins labelled with TMT and 15 proteins that were not labelled (data not shown), confirming that 98.5% of the peptides identified were successfully labelled. The mass spectrometry analysis of the fractionated digest identified 7352 human peptides (FDR>1%) corresponding to 1931 proteins. DENV proteins were not identified in any of the samples. We compared the proteome of two healthy controls, four DF patients (without evidence of plasma leakage) and four DHF patients (with plasma leakage). One healthy control (H1) was considered the reference sample and fold change of protein expression in all the samples were calculated to this reference sample. The mean fold change was calculated for the groups (healthy, DF and DHF) and a t-test was used to find a significant association among groups. Proteins with a significant fold change (>1.5) and a p<0.05 were considered differentially expressed during group comparisons.

### Proteins differentially regulated in dengue infected PBMCs compared to healthy controls

We identified 160 differentially regulated proteins, including 94 upregulated and 66 down-regulated proteins, with a fold change of >1.5 and p<0.05, when comparing the eight DENV-infected samples with healthy control samples (Figure 1a). These proteins were further analyzed to identify the main gene ontology (GO) pathways involved in DENV-infected PBMC, to determine the number of proteins involved and the fold enrichment (Figure 2). GO enrichment analysis revealed 197 (FDR ≤ 0.05) pathways up-regulated and 122 (FDR ≤ 0.05) pathways down-regulated in DENV infected PBMCs. Response to cytokines, cytokine-mediated signaling pathways, type 1 interferon signaling and defense response to virus were upregulated as expected in DENV-infected PBMCs during early infection. Most upregulated proteins were predicted to be localized in the MHC class I peptide loading complex and postsynaptic endocytic zone in the cells. All proteins differentially regulated in DENV-infected samples are listed in Supplementary Table 1. The heatmap and the pathway analysis is shown in figure 3.

**Figure 1:**
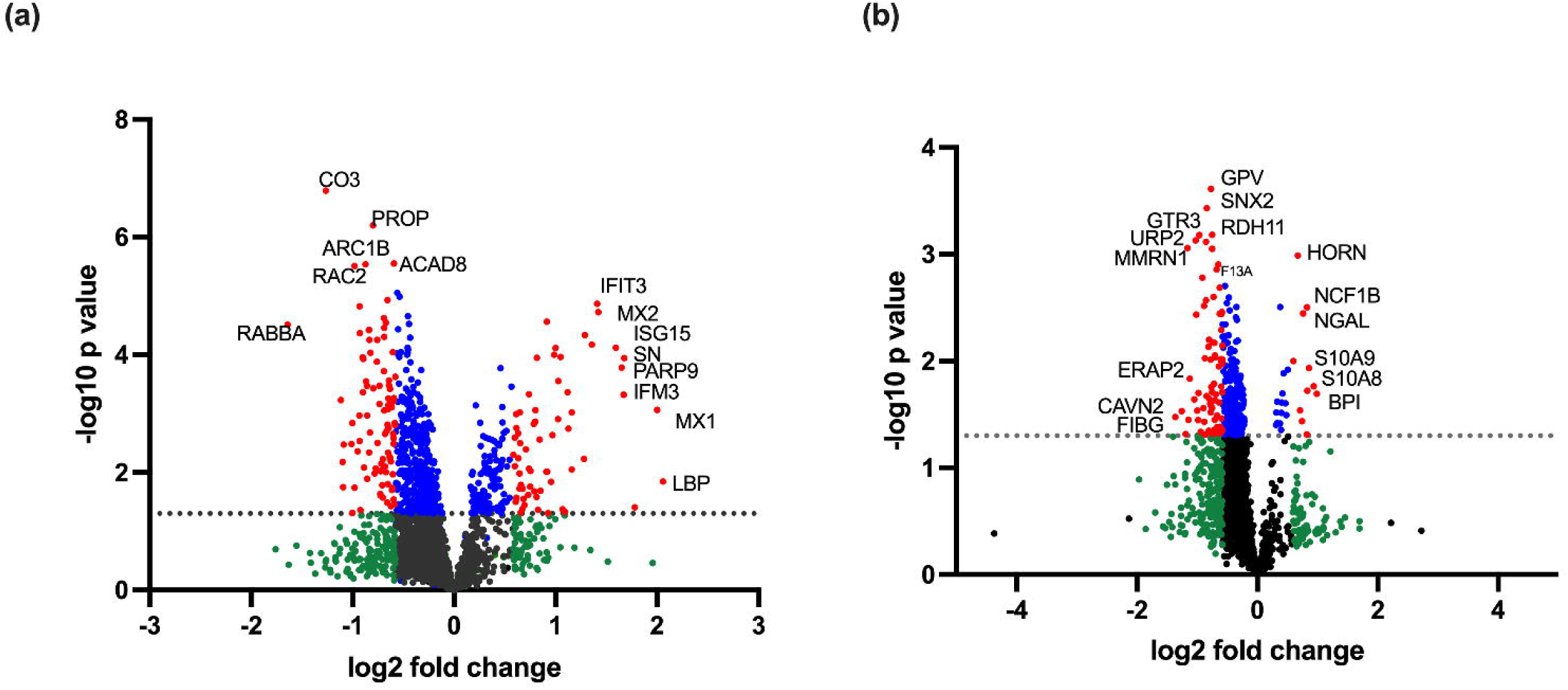
The volcano plots represent the fold change (log_2_ fold change) and the p value following t-test (-log_10_ p value) of the expression level of each protein detected in PBMC lysates as analysed by TMT mass spectrometry. Black represents proteins with no difference in expression; green represents a fold change difference of > 1.5 without a statistically significant p value (p>0.05); blue represents a fold change of <1.5 with a statistically significant p value (p<0.05); red represents proteins with a fold change of > 1.5 and a statistically significant difference (p<0.05) among groups. Proteins having high -log_10_ p value and log_2_ fold change were labelled with their names. (a) Mean values of each protein expressed in the dengue-infected 8 samples were compared to the mean expression values of the 2 healthy controls. 210 proteins fell into the green category, 502 into the blue and 160 into the red category. (b) Mean values of each protein expressed in the 4 DHF samples were compared to those of the 4 DF samples. 278 were in the green, 232 in the blue and 90 in the red category.

**Figure 2:**
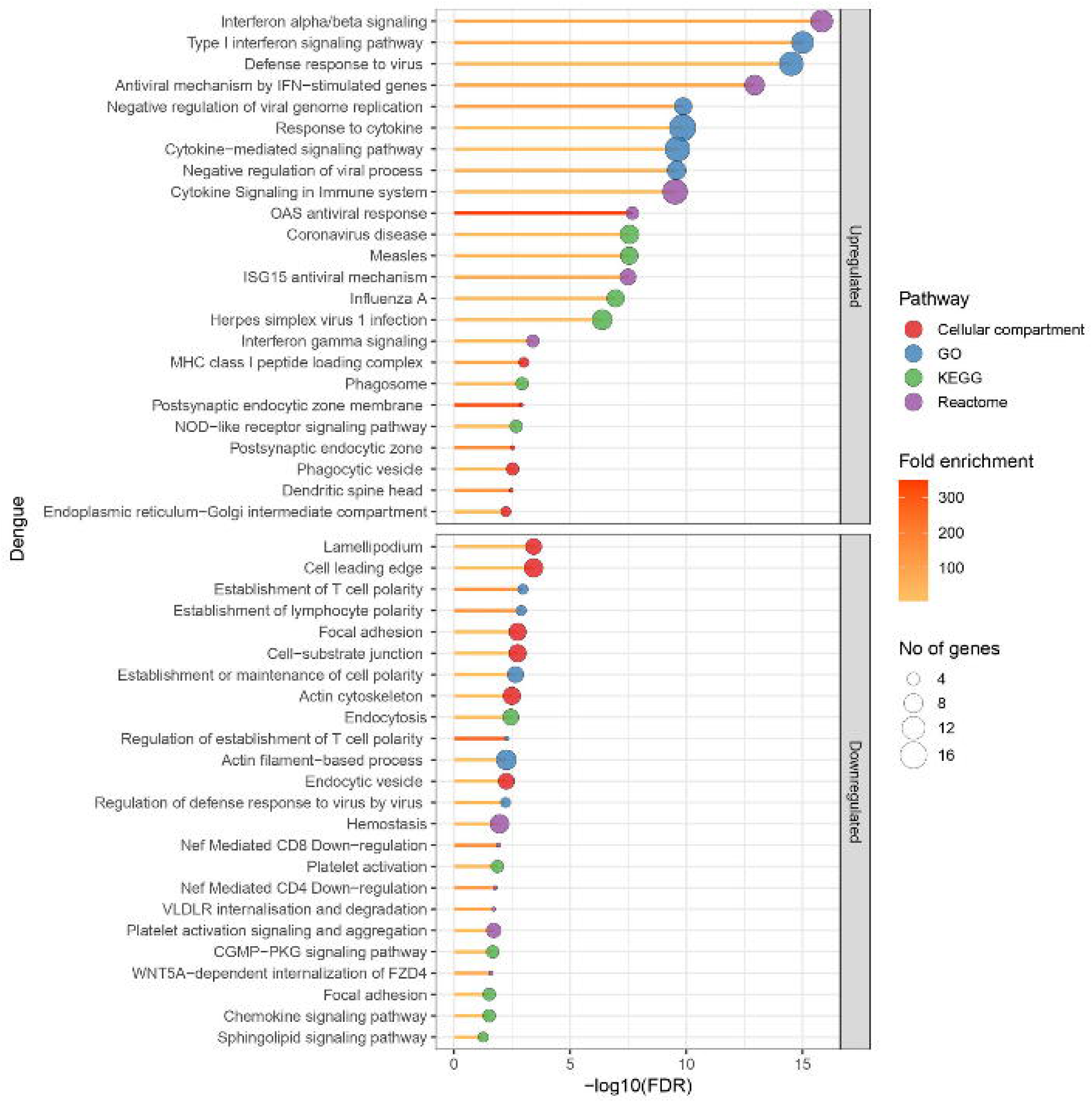
Gene ontology (GO) pathways identified to be differentially regulated in the dengue-infected PBMC lysates compared to healthy controls. Two healthy control PBMC lysates and eight dengue-infected lysates were subjected to TMT tag mass spectrometry. Mean protein expression levels among healthy samples were compared to those of dengue-infected samples. Proteins with a fold change of >1.5 and a statistically significant difference in dengue-infected lysates compared to healthy controls were subjected to STRING pathway analysis. Main pathways differentially regulated in dengue-infected lysates are given in the figure. The size of the blue circle corresponds to the number of proteins involved in each pathway and the yellow-red colour code corresponds to the fold enrichment.

**Figure 3:**
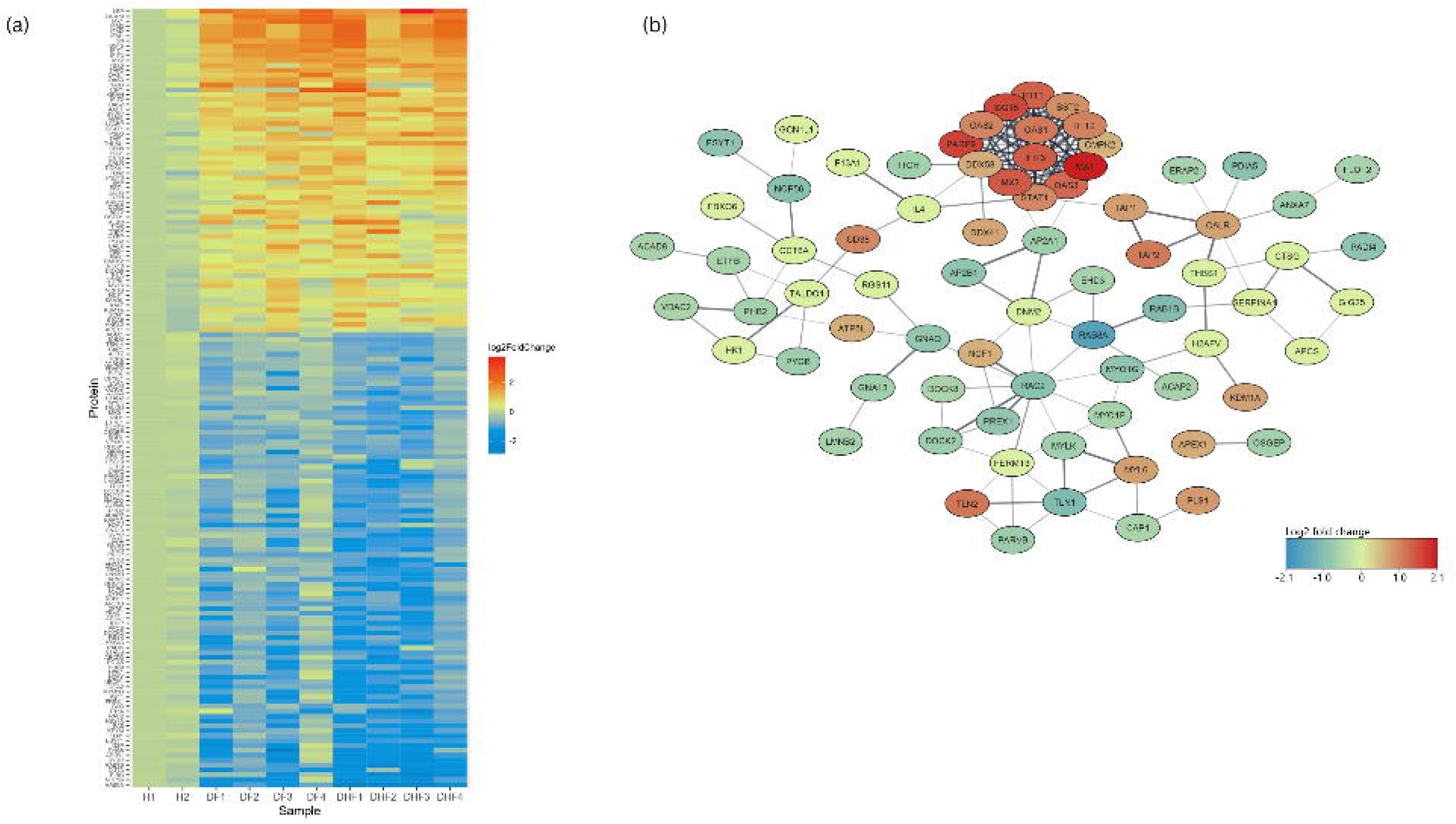
Differentially regulated proteins in dengue-infected PBMC lysates compared to the healthy controls. (a) The 160 proteins expressed to a significantly lower or higher level in the DV-infected PBMC lysates (DF1-4 and DHF 1-4) compared to the healthy control sample 1 (H1) are shown in the heat map. The red colour indicates a higher expression level while the blue colour indicates a lower expression level in dengue samples compared to H1. (b) The protein network of the differentially regulated proteins identified in the DV-infected samples by STRING software.

### Proteins differentially regulated in DHF compared to DF

Ninety proteins were differentially expressed in DHF compared to DF PBMC with a fold change of >1.5 and p<0.05 (Figure 1b). Twelve proteins had increased and 78 had decreased expression levels in DHF lysates (Figure 4). GO enrichment revealed a total of 142 upregulated and 150 downregulated pathways. Highly enriched pathways in DHF were involved in negative regulation of macrophage activation and interleukin-8 production, as well as positive regulation of the p38MAPK cascade and ERBB signaling. Most upregulated proteins were localized in phagolysosomes, lysosomes and the NADPH oxidase complex (Figure 5). Down-regulated proteins were mainly involved in coagulation which included platelet activation and aggregation. Many downregulated proteins were localized at cell-substrate junctions, focal adhesion and anchoring junctions in the cells. All differentially expressed proteins are listed in Supplementary Table 2. Selected proteins of interest are shown in Figure 6.

**Figure 4:**
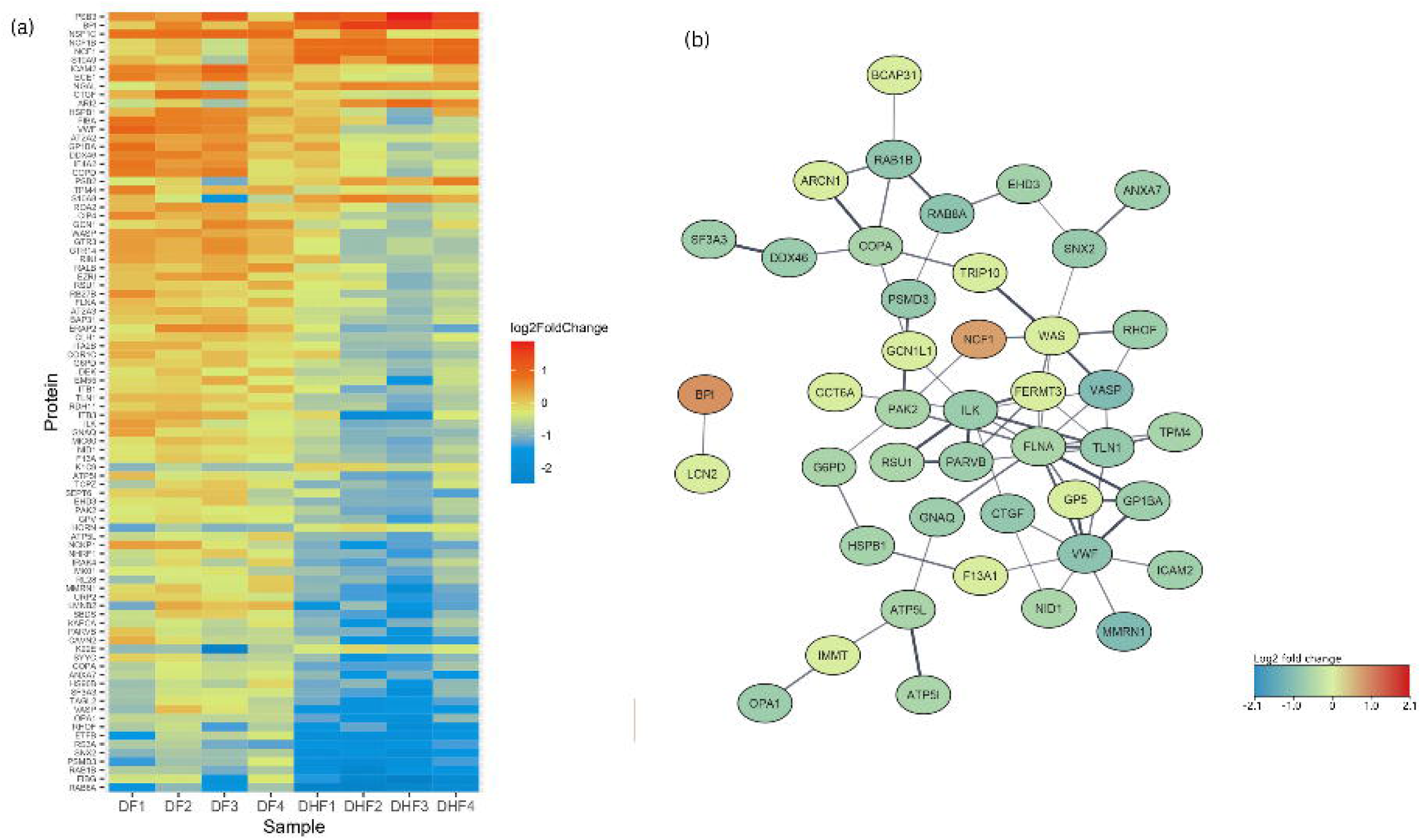
Differentially regulated proteins in dengue fever (DF) patient PBMC lysates compared to dengue haemorrhagic fever (DHF). (a) The 90 proteins that were expressed at significantly lower or higher levels in DF compared to DHF PBMC lysates (DF 1-4 vs DHF 1-4) after normalization to healthy control sample 1 (H1), are shown in a heat map. The red colour indicates a higher expression level while the blue colour indicates a lower expression level among DHF samples compared to DF sample 1. (b) Protein network of the differentially regulated proteins identified in the DHF samples compared to DF samples as analyzed by STRING software.

**Figure 5:**
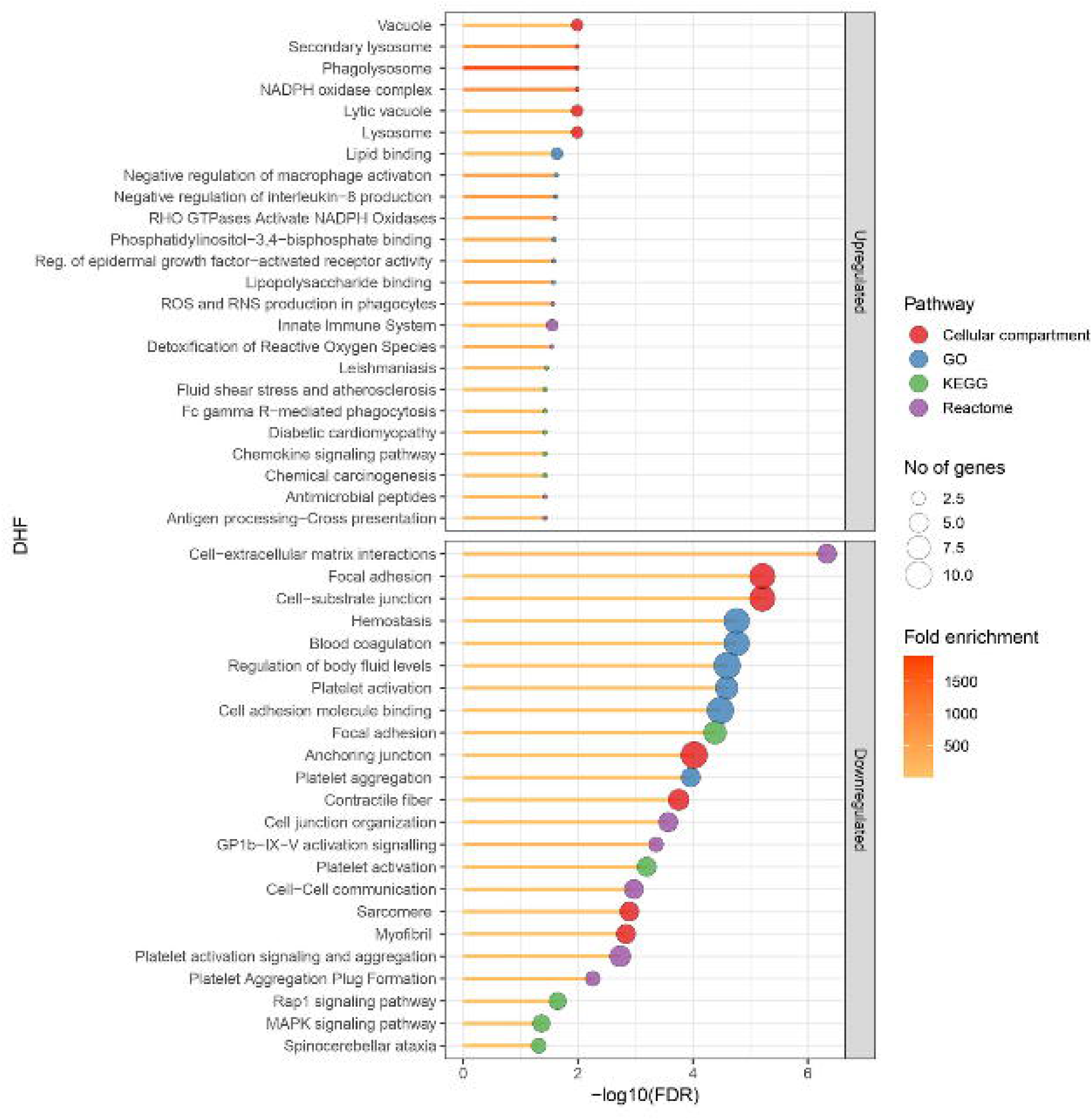
Gene ontology (GO) pathways identified to be differentially regulated in the dengue haemorrhagic fever (DHF) patient PBMC lysates compared to dengue fever (DF) samples. Four DF patient PBMC lysates and four DHF patient lysates were subjected to TMT tag mass spectrometry. Mean protein expression levels of DF samples were compared to those of the DHF samples. Proteins with a fold change of >1.5 and a statistically significant difference in DHF patient lysates compared to DF samples were subjected to STRING pathway analysis. The main differentially regulated pathways are shown in the figure. The size of the blue circle corresponds to the number of proteins involved in each pathway and the yellow-red colour code corresponds to the fold enrichment.

**Figure 6:**
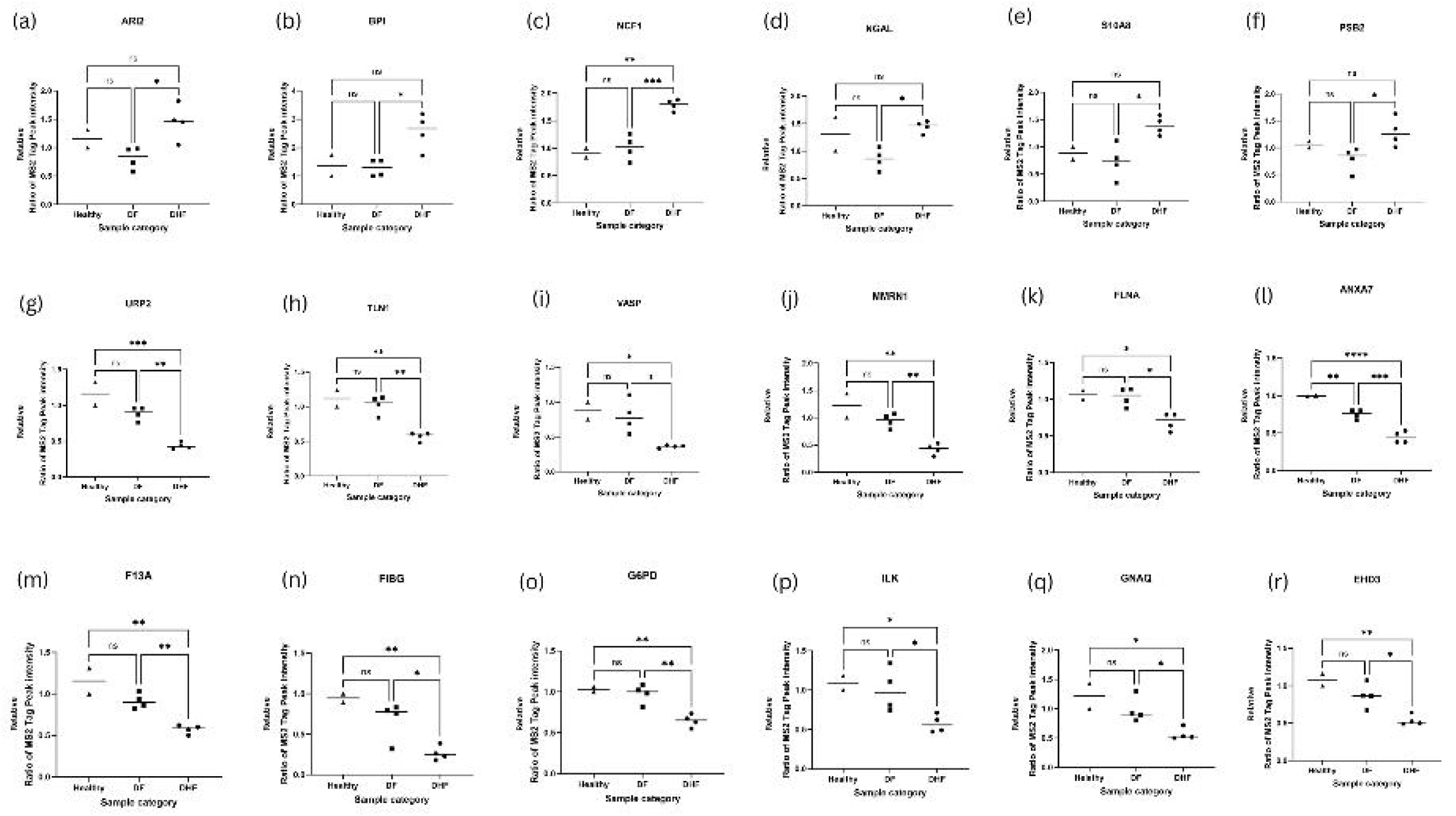
Selected proteins of interest identified by TMT tag mass spectrometry in PBMC lysates of healthy (n=2), DF (n=4) and DHF (n=4) patients. Figure represents individual selected proteins expressed as a ratio compared to the H1 sample, statistically different among healthy, DF and DHF groups analysed by ANOVA. (a) - (f) show proteins found at higher levels in DHF compared to DF lysates. (g) - (r) show proteins with a reduced expression level in DHF compared to DF samples. *p<0.05, **p<0.01, **p<0.001, ***p<0.0001

## Discussion

DHF can be fatal if not managed appropriately. Due to a lack of markers that could accurately predict development of plasma leakage, patients with significant thrombocytopaenia are admitted to hospitals and monitored closely. This results in significant overcrowding of hospitals in many endemic countries with overburdening of staff. Therefore, identification of biomarkers that could accurately predict disease severity in dengue fever patients early during the course of disease has been at the heart of much ongoing research. Variable and non-reproducible results have been a problem in utilizing for example inflammatory mediators as potential markers. Previous transcriptomic analysis of patient PBMC identified increased expression of RNA transcripts involved in mitochondrial processes and neutrophil associated enzymes (MO, ELANE) in severe dengue (9,10). Nikolayeva *et al.* identified an 18-gene panel in PBMC for detection of severe dengue on admission to hospital (12). Although transcriptomic data provide useful information, identification of proteins provides a more robust indication of cellular activity and proteins are also useful in developing cost-effective lateral flow tests. Platelet activation and sequestration of histones were seen in platelets of dengue-infected samples in a proteomic analysis (13). This proteomic analysis revealed important observations on the role of platelets in dengue infection but it was not designed to find an association with disease severity. Currently, there are no approved biomarkers that could be used to triage patients in early disease. Here we report for the first-time which proteins are differentially expressed in PBMC of DHF patients compared to those with non-severe disease. This study was performed using samples of patients during the early disease stage, before they developed plasma leakage. The proteins identified here could therefore, once validated in a larger cohort, potentially be useful biomarkers for detecting DHF early in the disease course, in addition to shedding light on pathways potentially contributing to plasma leakage.

We did not identify any DENV proteins in the PBMC samples. This is not surprising due to the low levels of virus proteins expected in the infected cells. Dengue-infected PBMC showed increased expression levels of proteins involved in interferon-signalling compared to healthy control samples as observed previously in transcriptomic analysis of DENV-infected PBMCs (9,10). Type I interferons elicit antiviral responses to protect the host through many interferon-stimulated genes (ISGs) and the ISG15 protein plays a central role in this defence response (14). ISG15 was highly expressed in DENV-infected PBMC compared to healthy controls indicating the type I interferon-driven antiviral response. However, ISG15 was not differently expressed in DHF compared to DF which confirms similar findings from previously published transcriptomic data (10). In addition, proteins involved in inflammatory signalling which includes cytokine activation and proteins localised in the phagocytic vesicles were also expressed at higher levels in DENV infected PBMC to combat the virus. Proteins involved in the cytoskeleton, cell-substrate junction and actin-filament based processes were down-regulated in dengue infected PBMC. This reduced expression of proteins of the cytoskeleton and cell adhesion was more pronounced in patients who developed DHF compared to DF. The host cytoskeleton is known to play an important role in facilitating DENV entry to cells and viral replication (15). DHF PBMCs exhibited down-regulation of these processes suggesting a yet unidentified role in the development of vascular leakage possibly by leucocyte-endothelial cell interaction. Endothelial cells infected *in vitro* with DENV and treated with TNFα, showed reduced levels of moesin (a cytoskeletal protein) resulting in increased trans-endothelial permeability as reported previously (16). However, the exact mechanisms and cascade of events leading to increased vascular permeability is still unknown.

A negative effect on proteins involved in haemostasis, platelet activation, aggregation and signalling was observed in dengue-infected PBMC and this down regulation was more pronounced in the DHF samples compared to DF lysates. Thrombocytopaenia is known to be more pronounced in DHF and it predicts a worse outcome (17). Many proteins such as TLN1 (18), FLNA (19), VWF (20), important in haemostasis and platelet function were down-regulated in DHF PBMCs during the early disease course suggesting that bleeding manifestations seen in DHF could partly be due to these suppressed pathways. In addition, FXIIIA (factor 13 subunit A), which circulates in plasma was significantly reduced in PBMCs of patients who progressed to DHF. FXIII has two subunits, of which the A subunit is secreted from cells of myeloid lineage and platelets (21). Subunit B is secreted from the hepatocytes (21). Although the main function of FXIII is to stabilise the clot, many other functions of this coagulation factor in monocytes are recognized mainly related to phagocytosis and interaction with the cytoskeleton (22). Plasma levels of FXIIIA can be measured and it could be a useful marker to identify development of DHF in future studies.

Two important observations were the increased activation of oxidative stress-related pathways and activation of p38 mitogen-activated protein (MAP) kinase signalling in DHF samples. MAP kinases are important intracellular kinases involved in regulation of cellular proliferation, differentiation, and apoptosis in addition to their role in cellular inflammation (23). p38 MAPK activation results in production of pro-inflammatory cytokines such as IL-1β, IL-6, TNFα (24), and p38 has been reported to regulate TNFα gene transcription (25). *In vitro* studies have shown that DENV-infected endothelial cells express increased amounts of p38 MAP kinases and it is postulated that p38 signalling increases levels of pro-inflammatory mediators such as IL-8 (26). p38 MAPK was also activated in endothelial cells by DENV NS1 protein resulting in increased endothelial permeability (7) inhibition of p38 signalling *in vivo,* reduced inflammatory response, vascular leak and improved survival of mice (27). Cross-talk between dengue-infected immune cells and the endothelium could be important in disruption of endothelial integrity and development of vascular leak. Thus, findings in the PBMC of patients who subsequently developed DHF indicate a p38 MAPK-driven response potentially contributing to severe disease. In addition, proteins involved in this pathway could be developed into early biomarkers predicting severe disease.

Oxidative stress is known to play a role in DENV pathogenesis (28). Reactive oxygen species (ROS) activate inflammatory pathways in immune cells (28) and induce vascular permeability in endothelial cells (29). Markers of oxidative stress such as malondialdehyde are elevated in DHF (30). Upregulated proteins in DHF samples implicated in oxidative stress could be useful as markers of fluid leakage following validation in larger cohorts.

Neutrophil associated transcripts were abundant in previous transcriptomic analysis of PBMCs of patients who developed dengue shock syndrome (11). Consistent with this study, we identified BPI (bactericidal permeability increasing) protein to be high in the DHF patient PBMCs. The role of neutrophils in dengue infection is not clear. Studies have shown that dengue infection results in *in vivo* neutrophil activation and neutrophil extracellular trap formation (31). The authors hypothesized that a possible role of neutrophil extracellular traps may be in mediating vascular leakage by disrupting the endothelial barrier (31). NGAL (Neutrophil Gelatinase associated Lipocalin) is a protein initially purified from neutrophils, but expressed in many organs and cells. A previous study showed a higher serum level of NGAL in dengue-infected individuals compared to healthy controls (32). Levels correlated with white blood cells and platelets. However, a relationship to disease severity was not investigated in this study. NGAL is one of the few proteins highly expressed in PBMCs of patients who progressed to DHF compared to DF. Therefore, these proteins could be potential markers to identify DHF early in the disease course in future validation studies.

In conclusion, we have for the first time explored the proteome of PBMC from DENV-infected patients during early infection to identify potential markers predicting DHF as well as pathways implicated in vascular leakage. Our study was aimed at identifying proteins from PBMC lysates to gain a better understanding how processes within these cells may cause, reflect and perhaps also predict the further course of disease. To develop promising proteins into useful biomarkers, a next step will be to look at the secretion of selected proteins into the bloodstream and validate them in larger cohorts as potential biomarkers to predict the risk of developing DHF during early stages of dengue infection.

## Materials and methods

### Study setting and patient recruitment

A total of 25 dengue patients fulfilled inclusion criteria and they were recruited from the National Institute of Infectious Diseases, Sri Lanka. Of these patients, 17 had DF and 8 patients progressed to DHF in the subsequent days. The study participants were recruited from the outpatient department and the wards. The criteria for inclusion in the study were; patients with a febrile illness suggestive of dengue fever lasting less than 72 hours. Patients less than 18 years of age were excluded due to complexities involved in obtaining consent. In addition, pregnant patients were also excluded due to diverse factors affecting the course of illness. Any patient with evidence of fluid leakage on recruitment was excluded from the study to facilitate recruitment of patients who were at an early disease stage. The SD Bioline dengue rapid NS1 kit (Abbott) was used to identify patients with dengue fever on recruitment.

The study was explained to the participants and written consent was obtained at recruitment. After obtaining their consent, venous blood samples were taken at admission. A 5 ml blood sample was collected into a 15 ml heparinised container for PBMC isolation and 2 ml of blood was collected in a plain tube for separating serum by centrifugation at 2000rpm. All participants had the right to withdraw from the research during the entire study period. Clinical data were obtained from the bed-head tickets and samples were initially processed in the Centre for Dengue Research at University of Sri Jayewardenepura and final proteomics experiments were carried out in the Oxford Glycobiology Institute, University of Oxford. Fifteen healthy volunteers were recruited to generate samples to be used as controls from Sri Lanka during the study period. The study was approved by the Oxford Tropical Research Ethics Committee (OxTREC 16/18) and the Ethics Review committee (ERC 01/18), University of Sri Jayewardenepura, Sri Lanka prior to the commencement of recruitment.

### Measurement of viral load in serum

RNA extraction of serum samples was performed using the QiaAmp viral RNA mini kit (Qiagen) using 140 μL of serum according to the manufacturer’s instructions. The extracted RNA was reverse transcribed to cDNA using the high-capacity cDNA reverse transcription kit (ThermoFisher Scientific). Following reverse transcription, 2 μL of cDNA was used in a 20 μL qRT-PCR reaction which comprised 10 μL Taqman multiplex mastermix (ThermoFisher Scientific), 900 nM of each primer and 250 nM of each probe to amplify and measure DENV copies. The standard curve was generated by a mixture of gBlock DNA fragments similar to the sequences amplified in the PCR reaction and the viral load in the sample was calculated using the standard curve.

The DENV serotype was identified and the viral load was measured by a multiplex qRT-PCR using four different Taqman primers and dual labelled probes for the four serotypes. All probes were labelled with the QSY quencher. DENV targets and primers used for quantifying viral load by qRT-PCR are given in Table 2.

**Table 2:**
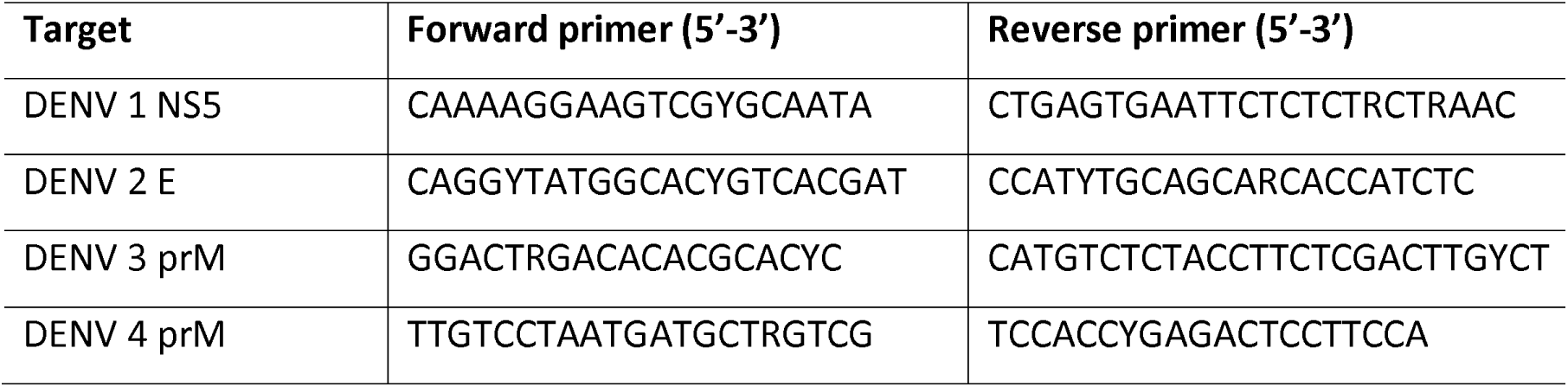
Target genes and the primer sequences used for qRT-PCR of DENV RNA.

### PBMC isolation

The PBMC were isolated by a gradient centrifugation method. Whole blood (5 ml) was collected in a heparinised 15 ml falcon tube and transported to the laboratory at room temperature for PBMC isolation within 4 hours. Blood was diluted 1:1 in sterile phosphate buffered saline (PBS) and overlayed slowly onto 5 ml of Lymphoprep (STEMCELL Technologies). The sample was centrifuged at 2000 rpm (900g) at room temperature for 20 min, with the brake off. The PBMC layer was aspirated using a sterile Pasteur pipette and transferred to a new tube. Ice cold PBS was added to the cells and they were centrifuged at 1500 rpm (500g), for 10 min at 4 °C, with the brake on low. The supernatant was discarded and the cell pellet was resuspended in 1x red blood cell lysis buffer (BioLegend) in deionized water and incubated at 4°C for 5 min to remove any red cells. The lysis buffer was removed by adding 14 ml PBS and centrifuged at 960 rpm (200g) for 10 min at 4°C, with the brake on high. Cells were washed once in PBS and lysed to generate cell lysates for mass spectrometry.

### Tandem mass tagging

PBMC cell lysates were produced by resuspending the cell pellets in RIPA buffer, supplemented with protease inhibitor (Roche), phosphatase inhibitor (Sigma) and incubating at 4°C for 20 min. The lysates were centrifuged at 12,000g for 10 min and the supernatant collected for experiments. The lysates were subjected to quantification of protein levels by the Pierce BCA protein assay kit (ThermoFisher Scientific). The four DF and four DHF samples with the highest protein concentrations present were selected for tandem mass tagging (TMT). The protein amount was adjusted to 7.5 μg for each sample and was run for 5 min (approximately 5 mm) into a 4-12% Bis-Tris 10-well SDS-PAGE gel using the MOPS buffer system (ThermoFisher Scientific). The gel was stained with InstantBlue protein stain (Exedeon) and the bands were excised from the gel with a scalpel and transferred to fresh tubes. In-gel trypsin digestion was carried out as previously described(33) using sequencing grade porcine trypsin (Promega). Further normalisation was performed by determining the peptide concentrations of the samples using the Pierce quantitative colorimetric peptide assay (ThermoFisher Scientific). Labelling was carried out using the TMT10plex isobaric labelling kit (ThermoFisher Scientific) according to the manufacturer’s recommended protocol. The labels for the 10 plex kit were used with the following tags in parentheses: two healthy donors (126, 127N), four DF (127C, 128N, 128C, 129N) and four DHF (129C, 130N, 130C, 131) patients where the number refers to the mass of the reporter in Daltons and if two tags have the same mass then N indicates if the nitrogen in the mass reporter is a heavy stable isotope and C indicates if only carbons are heavy labelled in the mass reporter. Each TMT 10-plex tag (19 μl) was mixed with 50 μl of each normalised peptide sample and then 60 μl of each of these 10 samples were pooled. This pooled sample (600 μl) was dried in a Concentrator Plus centrifugal evaporator (Eppendorf) and then resuspended in 300 μL of 0.1%v/v TFA. As a check for labelling, 5 μl of this unfractionated sample was run by liquid chromatography-mass spectrometry (LC-MS) and the data was searched on the Mascot software as described below. After confirming reliable labelling efficiency, the remaining 295 μL of pooled sample was fractionated into 11 fractions using the Pierce high pH reversed-phase peptide fractionation kit (ThermoFisher Scientific) according to the manufacturer’s recommended protocol. These 11 fractions were run by LC-MS (25 μl for fractions 1-3 and 15 μl for all other fractions) and the data was searched on the PEAKS Studio X software as described below.

### Liquid chromatography

The 11 TMT labelled peptide fractions were separated on a Dionex Ultimate 3000 nano UHPLC system (ThermoFisher Scientific). A nano analytical C18 reversed phase column (PepMap) was used (column dimensions 75 μm diameter, 50 cm length, 2 μm particle size) (ThermoFisher Scientific) with a flow rate of 250 nL/min at 45°C. The mobile phase used was solvent A: 0.1% v/v formic acid in LC-MS grade water, and solvent B: 0.1% v/v formic acid in 80% v/v acetonitrile. The gradient was used to run the peptides for the TMT labelling efficiency check as previously described (34). A longer 3 hour gradient was used to separate peptides from the 11 fractions as follows: 2% B (0-6.6 min), 2-35% B (6.6-150 min), 35-60% B (150-185 min), 60-95% B (185-186 min), 95% B (186-194 min), 95-2% B (194-195 min) and 2% B (195-210.1 min) for column equilibration.

### Mass spectrometry

Peptides from the nano LC were analysed on a benchtop Q Exactive hybrid quadrupole-Orbitrap mass spectrometer using the Nanospray Flex ion source (ThermoFisher Scientific). Prior to data acquisition, the mass spectrometer was calibrated for mass accuracy according to the manufacturer’s recommendations using a positive ion calibration solution injected at 5 µl/min into a heated electrospray ionisation (HESI) probe (ThermoFisher Scientific). The conditions for data dependent acquisition (DDA) were as follows: chromatographic peak width was set at 20 s and the Full MS conditions were with a resolution of 70,000, AGC target of 3e6, maximum IT (injection time) of 60 ms, scan range of 375 to 1500 m/z. The dd-MS2 conditions were with a resolution of 35,000, AGC target of 1e5, maximum IT of 60 ms, loop count of 10 (i.e. Top 10), isolation window of 2.0 m/z, fixed first mass of 120.0 m/z and normalised collision energy (NCE) was 35 in a high-energy collision dissociation (HCD) cell. The data-dependent (dd) settings were with minimum intensity threshold of 3.3e4 ions, charge exclusion: unassigned, 1, 8, >8, peptide match: preferred, dynamic exclusion: 30 s.

### Protein identification

The acquired *.raw file for the 5 μl labelling check was converted to a Mascot generic file (*.mgf) using MSConvertGUI 64-bit (ProteoWizard). A peak-picking filter was set with MS levels 1-2. The Mascot server software (Matrix Science, London) was used to search the *.mgf file against the SwissProt database. The search parameters in Mascot were set as follows: MS/MS Ion Search with trypsin as the protease. Carbamidomethyl (C) and Oxidation (M) were set as variable modifications. Taxonomy was set to *Homo sapiens* (human).

Peptide mass tolerance and fragment mass tolerance were both set as ± 10 ppm and the peptide charge was set as +2, +3 and +4. Data format and instrument were set as Mascot generic and Q Exactive, respectively. A decoy database was searched with the false discovery rate (FDR) adjusted to 1%. Searches were carried out with both no fixed modification and TMT as a fixed modification to calculate the labelling efficiency. The percentage of peptides labelled with TMT was calculated as follows: (Number of proteins identified with TMT 10plex ÷ Total number of peptides identified) x 100.

Once a high percentage of labelling was determined, the high pH reversed-phase fractionation and LC-MS runs of each fraction was carried out. The 11 *.raw files for each fraction were searched on the PEAKS Studio (version X) software against the Uniprot_SwissProt database. FDR was set at <1% and precursor mass tolerance to 10 ppm. Trypsin was set as the protease, Oribtrap as the instrument, CID\HCD as the fragmentation and MS2 as the reporter ion type. TMT was set as a fixed modification. Both Carbamidomethyl (C) and Oxidation (M) were set as variable modifications. Taxonomy was set to both human and dengue virus.

### Proteomic data analysis

All human proteins identified in the samples were selected for data analysis. The average value of the MS2 tag peak intensity was calculated for proteins with multiple peptides. The mean MS2 tag peak intensity ratio for each peptide was calculated for the healthy, DF and DHF samples. The H1 healthy sample was used as the reference sample to calculate this ratio as the relative MS2 tag peak intensity for each sample. The fold change for each protein was calculated for dengue-infected samples (DF and DHF both) compared to healthy controls (H1 and H2) and DF (DF 1-4) compared to DHF (DHF 1-4). A t-test was performed among samples to identify a statistically significant difference of protein expression between healthy controls and dengue-infected samples. A similar analysis was performed for DF and DHF. Proteins with a fold difference exceeding 1.5 and p<0.05 were selected to identify biological pathways associated with the proteins using STRING database. Figures were generated using GraphPad Prism 9.0 version and R studio with R v4.1.2. Protein to protein interaction (PPI) networks were created using Cytoscape v3.9.1. Gene ontology enrichment analysis was conducted by calculating probabilities and fold enrichment values from observed differently expressed proteins. Gene Ontology (GO) database (35) was used to enrich biological processes, molecular function and cellular components while Kyoto Encyclopedia of Genes and Genomes (KEGG) and REACTOME databases were used to enrich pathways (36,37). Fold enrichment and false discovery rate (FDR) were calculated for each pathway. Fold enrichment measures how drastically the genes of a certain pathway is overrepresented, which was calculated by the percentage of differently expressed proteins belonging to a pathway divided by the corresponding percentage in the background. FDR was calculated based on nominal P-value from the hypergeometric test which represents the statistical probability of each enrichment. A cut-off of 0.05 was used on the FDR to filter enriched pathways. Calculations were done using R\GO.db and R\Gostats packages, and visualizations were done using R\ ggplot2.

## Data Availability

All data produced in the present study are available upon reasonable request to the authors

## Acknowledgements

We acknowledge staff of National Institute of Infectious Diseases for their valuable support in conducting the study and Dr Parami Rajapakse for her assistance in formatting figures for the paper.

