## Supplementary table 1 for "Proteomics analysis of peripheral blood monocytes from patients in early dengue infection reveals potential markers of subsequent fluid leakage"

**Supplementary Table 1:** Differentially expressed proteins in dengue-infected cell lysates compared to healthy controls

| **Accession** | **Protein names** | **log_2_ fold change** | **-log_10_ p value** |
| --- | --- | --- | --- |
| RAB8A | Ras-related protein Rab-8A (EC 3.6.5.2) (Oncogene c-mel) | -1.6408 | 4.51041 |
| SYDC | Aspartate--tRNA ligase, cytoplasmic (EC 6.1.1.12) (Aspartyl-tRNA synthetase) (AspRS) (Cell proliferation-inducing gene 40 protein) | -1.1148 | 3.23091 |
| CUTA | Protein CutA (Acetylcholinesterase-associated protein) (Brain acetylcholinesterase putative membrane anchor) | -1.0991 | 2.17585 |
| TLN1 | Talin-1 | -1.0935 | 1.74936 |
| RAB1B | Ras-related protein Rab-1B (EC 3.6.5.2) | -1.087 | 2.47392 |
| FIBG | Fibrinogen gamma chain | -1.0096 | 2.48404 |
| NOP56 | Nucleolar protein 56 (Nucleolar protein 5A) | -1.0028 | 1.31267 |
| DHB8 | (3R)-3-hydroxyacyl-CoA dehydrogenase (EC 1.1.1.n12) (17-beta-hydroxysteroid dehydrogenase 8) (17-beta-HSD 8) (HSD17B8) (3-ketoacyl-[acyl-carrier-protein] reductase alpha subunit) (KAR alpha subunit) (3-oxoacyl-[acyl-carrier-protein] reductase) (Estradiol 17-beta-dehydrogenase 8) (EC 1.1.1.62) (Protein Ke6) (Ke6) (Short chain dehydrogenase/reductase family 30C member 1) (Testosterone 17-beta-dehydrogenase 8) (EC 1.1.1.239) | -1.0025 | 2.83755 |
| PDIA5 | Protein disulfide-isomerase A5 (EC 5.3.4.1) (Protein disulfide isomerase-related protein) | -0.9802 | 1.73983 |
| RAC2 | Ras-related C3 botulinum toxin substrate 2 (GX) (Small G protein) (p21-Rac2) | -0.9802 | 5.50963 |
| PADI4 | Protein-arginine deiminase type-4 (EC 3.5.3.15) (HL-60 PAD) (Peptidylarginine deiminase IV) (Protein-arginine deiminase type IV) | -0.9498 | 2.35834 |
| ESYT1 | Extended synaptotagmin-1 (E-Syt1) (Membrane-bound C2 domain-containing protein) | -0.9334 | 2.96682 |
| G3P | Glyceraldehyde-3-phosphate dehydrogenase (GAPDH) (EC 1.2.1.12) (Peptidyl-cysteine S-nitrosylase GAPDH) (EC 2.6.99.-) | -0.9315 | 2.53363 |
| IQGA2 | Ras GTPase-activating-like protein IQGAP2 | -0.931 | 4.82133 |
| RB27A | Ras-related protein Rab-27A (Rab-27) (EC 3.6.5.2) (GTP-binding protein Ram) | -0.9278 | 4.36408 |
| AP2B1 | AP-2 complex subunit beta (AP105B) (Adaptor protein complex AP-2 subunit beta) (Adaptor-related protein complex 2 subunit beta) (Beta-2-adaptin) (Beta-adaptin) (Clathrin assembly protein complex 2 beta large chain) (Plasma membrane adaptor HA2/AP2 adaptin beta subunit) | -0.9235 | 1.35898 |
| FA5 | Coagulation factor V (Activated protein C cofactor) (Proaccelerin, labile factor) [Cleaved into: Coagulation factor V heavy chain; Coagulation factor V light chain] | -0.8998 | 3.95782 |
| TRML1 | Trem-like transcript 1 protein (TLT-1) (Triggering receptor expressed on myeloid cells-like protein 1) | -0.8985 | 3.3608 |
| MYO1G | Unconventional myosin-Ig [Cleaved into: Minor histocompatibility antigen HA-2 (mHag HA-2)] | -0.8955 | 3.92792 |
| F13A | Coagulation factor XIII A chain (Coagulation factor XIIIa) (EC 2.3.2.13) (Protein-glutamine gamma-glutamyltransferase A chain) (Transglutaminase A chain) | -0.8864 | 2.08252 |
| ARC1B | Actin-related protein 2/3 complex subunit 1B (Arp2/3 complex 41 kDa subunit) (p41-ARC) | -0.8734 | 5.53607 |
| PREX1 | Phosphatidylinositol 3,4,5-trisphosphate-dependent Rac exchanger 1 protein (P-Rex1) (PtdIns(3,4,5)-dependent Rac exchanger 1) | -0.8689 | 3.54944 |
| UBCP1 | Ubiquitin-like domain-containing CTD phosphatase 1 (EC 3.1.3.16) (Nuclear proteasome inhibitor UBLCP1) | -0.8606 | 3.47106 |
| KPYM | Pyruvate kinase PKM (EC 2.7.1.40) (Cytosolic thyroid hormone-binding protein) (CTHBP) (Opa-interacting protein 3) (OIP-3) (Pyruvate kinase 2/3) (Pyruvate kinase muscle isozyme) (Threonine-protein kinase PKM2) (EC 2.7.11.1) (Thyroid hormone-binding protein 1) (THBP1) (Tumor M2-PK) (Tyrosine-protein kinase PKM2) (EC 2.7.10.2) (p58) | -0.8575 | 2.96647 |
| GNAQ | Guanine nucleotide-binding protein G(q) subunit alpha (Guanine nucleotide-binding protein alpha-q) | -0.857 | 1.89363 |
| ICAL | Calpastatin (Calpain inhibitor) (Sperm BS-17 component) | -0.857 | 1.89363 |
| STA5B | Signal transducer and activator of transcription 5B | -0.8402 | 4.41975 |
| RDH11 | Retinol dehydrogenase 11 (EC 1.1.1.300) (Androgen-regulated short-chain dehydrogenase/reductase 1) (HCV core-binding protein HCBP12) (Prostate short-chain dehydrogenase/reductase 1) (Retinal reductase 1) (RalR1) (Short chain dehydrogenase/reductase family 7C member 1) | -0.8383 | 4.4237 |
| TCPG | T-complex protein 1 subunit gamma (TCP-1-gamma) (CCT-gamma) (hTRiC5) | -0.8379 | 2.33815 |
| PPCS | Phosphopantothenate--cysteine ligase (EC 6.3.2.51) (Phosphopantothenoylcysteine synthetase) (PPC synthetase) | -0.8349 | 4.24995 |
| HXK1 | Hexokinase-1 (EC 2.7.1.1) (Brain form hexokinase) (Hexokinase type I) (HK I) (Hexokinase-A) | -0.8287 | 2.6769 |
| KS6A3 | Ribosomal protein S6 kinase alpha-3 (S6K-alpha-3) (EC 2.7.11.1) (90 kDa ribosomal protein S6 kinase 3) (p90-RSK 3) (p90RSK3) (Insulin-stimulated protein kinase 1) (ISPK-1) (MAP kinase-activated protein kinase 1b) (MAPK-activated protein kinase 1b) (MAPKAP kinase 1b) (MAPKAPK-1b) (Ribosomal S6 kinase 2) (RSK-2) (pp90RSK2) | -0.824 | 4.0348 |
| PROP | Properdin (Complement factor P) | -0.7991 | 6.19936 |
| IDHP | Isocitrate dehydrogenase [NADP], mitochondrial (IDH) (EC 1.1.1.42) (ICD-M) (IDP) (NADP(+)-specific ICDH) (Oxalosuccinate decarboxylase) | -0.7961 | 3.42847 |
| PUR9 | Bifunctional purine biosynthesis protein ATIC (AICAR transformylase/inosine monophosphate cyclohydrolase) (ATIC) [Cleaved into: Bifunctional purine biosynthesis protein ATIC, N-terminally processed] [Includes: Phosphoribosylaminoimidazolecarboxamide formyltransferase (EC 2.1.2.3) (5-aminoimidazole-4-carboxamide ribonucleotide formyltransferase) (AICAR formyltransferase) (AICAR transformylase); Inosine 5'-monophosphate cyclohydrolase (IMP cyclohydrolase) (EC 3.5.4.10) (IMP synthase) (Inosinicase)] | -0.7833 | 1.98293 |
| TBA4A | Tubulin alpha-4A chain (EC 3.6.5.-) (Alpha-tubulin 1) (Testis-specific alpha-tubulin) (Tubulin H2-alpha) (Tubulin alpha-1 chain) | -0.7722 | 2.06768 |
| KAPCA | cAMP-dependent protein kinase catalytic subunit alpha (PKA C-alpha) (EC 2.7.11.11) | -0.761 | 3.87938 |
| UN13D | Protein unc-13 homolog D (Munc13-4) | -0.7564 | 4.25309 |
| PARVB | Beta-parvin (Affixin) | -0.7544 | 2.50045 |
| ANXA7 | Annexin A7 (Annexin VII) (Annexin-7) (Synexin) | -0.7368 | 3.47118 |
| PYGB | Glycogen phosphorylase, brain form (EC 2.4.1.1) | -0.7357 | 3.16318 |
| DOCK2 | Dedicator of cytokinesis protein 2 | -0.7263 | 1.6351 |
| AP2A1 | AP-2 complex subunit alpha-1 (100 kDa coated vesicle protein A) (Adaptor protein complex AP-2 subunit alpha-1) (Adaptor-related protein complex 2 subunit alpha-1) (Alpha-adaptin A) (Alpha1-adaptin) (Clathrin assembly protein complex 2 alpha-A large chain) (Plasma membrane adaptor HA2/AP2 adaptin alpha A subunit) | -0.7255 | 2.0213 |
| RS11 | Small ribosomal subunit protein uS17 (40S ribosomal protein S11) | -0.7242 | 1.60808 |
| DPM1 | Dolichol-phosphate mannosyltransferase subunit 1 (EC 2.4.1.83) (Dolichol-phosphate mannose synthase subunit 1) (DPM synthase subunit 1) (Dolichyl-phosphate beta-D-mannosyltransferase subunit 1) (Mannose-P-dolichol synthase subunit 1) (MPD synthase subunit 1) | -0.7174 | 2.14828 |
| AHNK | Neuroblast differentiation-associated protein AHNAK (Desmoyokin) | -0.7077 | 2.04483 |
| RS4X | Small ribosomal subunit protein eS4, X isoform (40S ribosomal protein S4) (SCR10) (Single copy abundant mRNA protein) | -0.7069 | 1.78154 |
| DYN2 | Dynamin-2 (EC 3.6.5.5) | -0.7035 | 2.2243 |
| H2AY | Core histone macro-H2A.1 (Histone macroH2A1) (mH2A1) (Histone H2A.y) (H2A/y) (Medulloblastoma antigen MU-MB-50.205) | -0.7024 | 1.57177 |
| ITCH | E3 ubiquitin-protein ligase Itchy homolog (Itch) (EC 2.3.2.26) (Atrophin-1-interacting protein 4) (AIP4) (HECT-type E3 ubiquitin transferase Itchy homolog) (NFE2-associated polypeptide 1) (NAPP1) | -0.6923 | 3.72082 |
| VP13C | Intermembrane lipid transfer protein VPS13C (Vacuolar protein sorting-associated protein 13C) | -0.6922 | 4.62512 |
| RHG01 | Rho GTPase-activating protein 1 (CDC42 GTPase-activating protein) (GTPase-activating protein rhoGAP) (Rho-related small GTPase protein activator) (Rho-type GTPase-activating protein 1) (p50-RhoGAP) | -0.69 | 4.45492 |
| SORL | Sortilin-related receptor (Low-density lipoprotein receptor relative with 11 ligand-binding repeats) (LDLR relative with 11 ligand-binding repeats) (LR11) (SorLA-1) (Sorting protein-related receptor containing LDLR class A repeats) (SorLA) | -0.6876 | 4.30552 |
| MYLK | Myosin light chain kinase, smooth muscle (MLCK) (smMLCK) (EC 2.7.11.18) (Kinase-related protein) (KRP) (Telokin) [Cleaved into: Myosin light chain kinase, smooth muscle, deglutamylated form] | -0.6875 | 3.0512 |
| RHG15 | Rho GTPase-activating protein 15 (ArhGAP15) (Rho-type GTPase-activating protein 15) | -0.6801 | 2.09939 |
| GNA13 | Guanine nucleotide-binding protein subunit alpha-13 (G alpha-13) (G-protein subunit alpha-13) | -0.6794 | 2.65469 |
| OSGEP | tRNA N6-adenosine threonylcarbamoyltransferase (EC 2.3.1.234) (N6-L-threonylcarbamoyladenine synthase) (t(6)A synthase) (O-sialoglycoprotein endopeptidase) (hOSGEP) (t(6)A37 threonylcarbamoyladenosine biosynthesis protein OSGEP) (tRNA threonylcarbamoyladenosine biosynthesis protein OSGEP) | -0.6726 | 4.54695 |
| CPNE1 | Copine-1 (Chromobindin 17) (Copine I) | -0.6711 | 2.00788 |
| LEG10 | Galectin-10 (Gal-10) (Charcot-Leyden crystal protein) (CLC) (Eosinophil lysophospholipase) (Lysolecithin acylhydrolase) | -0.6633 | 2.70779 |
| ABCD3 | ATP-binding cassette sub-family D member 3 (EC 3.1.2.-) (EC 7.6.2.-) (70 kDa peroxisomal membrane protein) (PMP70) | -0.6588 | 1.48786 |
| CO3 | Complement C3 (C3 and PZP-like alpha-2-macroglobulin domain-containing protein 1) [Cleaved into: Complement C3 beta chain; C3-beta-c (C3bc); Complement C3 alpha chain; C3a anaphylatoxin; Acylation stimulating protein (ASP) (C3adesArg); Complement C3b alpha' chain; Complement C3c alpha' chain fragment 1; Complement C3dg fragment; Complement C3g fragment; Complement C3d fragment; Complement C3f fragment; Complement C3c alpha' chain fragment 2] | -0.6568 | 4.92904 |
| PHB2 | Prohibitin-2 (B-cell receptor-associated protein BAP37) (D-prohibitin) (Repressor of estrogen receptor activity) | -0.6551 | 2.8122 |
| EIF3L | Eukaryotic translation initiation factor 3 subunit L (eIF3l) (Eukaryotic translation initiation factor 3 subunit 6-interacting protein) (Eukaryotic translation initiation factor 3 subunit E-interacting protein) | -0.6519 | 1.8001 |
| URP2 | Fermitin family homolog 3 (Kindlin-3) (MIG2-like protein) (Unc-112-related protein 2) | -0.6504 | 3.07711 |
| ERAP2 | Endoplasmic reticulum aminopeptidase 2 (EC 3.4.11.-) (Leukocyte-derived arginine aminopeptidase) (L-RAP) | -0.6493 | 2.3333 |
| MYO1F | Unconventional myosin-If (Myosin-Ie) | -0.647 | 3.14186 |
| SPB10 | Serpin B10 (Bomapin) (Peptidase inhibitor 10) (PI-10) | -0.6464 | 3.25896 |
| HACD2 | Very-long-chain (3R)-3-hydroxyacyl-CoA dehydratase 2 (EC 4.2.1.134) (3-hydroxyacyl-CoA dehydratase 2) (HACD2) (Protein-tyrosine phosphatase-like member B) | -0.6456 | 2.7587 |
| MK01 | Mitogen-activated protein kinase 1 (MAP kinase 1) (MAPK 1) (EC 2.7.11.24) (ERT1) (Extracellular signal-regulated kinase 2) (ERK-2) (MAP kinase isoform p42) (p42-MAPK) (Mitogen-activated protein kinase 2) (MAP kinase 2) (MAPK 2) | -0.6419 | 3.48089 |
| K1H1 | Keratin, type I cuticular Ha1 (Hair keratin, type I Ha1) (Keratin-31) (K31) | -0.6409 | 3.56327 |
| LMNB2 | Lamin-B2 | -0.6345 | 2.26145 |
| SYYC | Tyrosine--tRNA ligase, cytoplasmic (EC 6.1.1.1) (Tyrosyl-tRNA synthetase) (TyrRS) [Cleaved into: Tyrosine--tRNA ligase, cytoplasmic, N-terminally processed] | -0.6314 | 2.23647 |
| FLOT2 | Flotillin-2 (Epidermal surface antigen) (ESA) (Membrane component chromosome 17 surface marker 1) | -0.6303 | 3.41973 |
| ACAP2 | Arf-GAP with coiled-coil, ANK repeat and PH domain-containing protein 2 (Centaurin-beta-2) (Cnt-b2) | -0.6294 | 2.24623 |
| SLFN5 | Schlafen family member 5 | -0.6241 | 2.18452 |
| EHD3 | EH domain-containing protein 3 (PAST homolog 3) | -0.6225 | 1.42641 |
| ACTZ | Alpha-centractin (Centractin) (ARP1) (Actin-RPV) (Centrosome-associated actin homolog) | -0.6169 | 3.17786 |
| TALDO | Transaldolase (EC 2.2.1.2) | -0.6138 | 3.1055 |
| AL4A1 | Delta-1-pyrroline-5-carboxylate dehydrogenase, mitochondrial (P5C dehydrogenase) (EC 1.2.1.88) (Aldehyde dehydrogenase family 4 member A1) (L-glutamate gamma-semialdehyde dehydrogenase) | -0.6107 | 1.70345 |
| H2AV | Histone H2A.V (H2A.F/Z) (H2A.Z variant histone 2) | -0.6102 | 1.73564 |
| ETFB | Electron transfer flavoprotein subunit beta (Beta-ETF) | -0.6037 | 1.96674 |
| 2AAA | Serine/threonine-protein phosphatase 2A 65 kDa regulatory subunit A alpha isoform (Medium tumor antigen-associated 61 kDa protein) (PP2A subunit A isoform PR65-alpha) (PP2A subunit A isoform R1-alpha) | -0.6023 | 4.04254 |
| TCPZ | T-complex protein 1 subunit zeta (TCP-1-zeta) (Acute morphine dependence-related protein 2) (CCT-zeta-1) (HTR3) (Tcp20) | -0.5999 | 3.20628 |
| AT2A3 | Sarcoplasmic/endoplasmic reticulum calcium ATPase 3 (SERCA3) (SR Ca(2+)-ATPase 3) (EC 7.2.2.10) (Calcium pump 3) | -0.5996 | 2.65293 |
| TSN14 | Tetraspanin-14 (Tspan-14) (DC-TM4F2) (Transmembrane 4 superfamily member 14) | -0.5963 | 2.58299 |
| GRAN | Grancalcin | -0.5952 | 2.48858 |
| DOCK8 | Dedicator of cytokinesis protein 8 | -0.5946 | 1.53804 |
| CAP1 | Adenylyl cyclase-associated protein 1 (CAP 1) | -0.5932 | 3.26462 |
| ACAD8 | Isobutyryl-CoA dehydrogenase, mitochondrial (IBDH) (EC 1.3.8.5) (Activator-recruited cofactor 42 kDa component) (ARC42) (Acyl-CoA dehydrogenase family member 8) (ACAD-8) | -0.5925 | 5.55364 |
| RS20 | Small ribosomal subunit protein uS10 (40S ribosomal protein S20) | -0.5869 | 1.45783 |
| ETHE1 | Persulfide dioxygenase ETHE1, mitochondrial (EC 1.13.11.18) (Ethylmalonic encephalopathy protein 1) (Hepatoma subtracted clone one protein) (Sulfur dioxygenase ETHE1) | -0.5854 | 2.83856 |
| VDAC2 | Voltage-dependent anion-selective channel protein 2 (VDAC-2) (hVDAC2) (Outer mitochondrial membrane protein porin 2) | -0.5804 | 2.80619 |
| STXB2 | Syntaxin-binding protein 2 (Protein unc-18 homolog 2) (Unc18-2) (Protein unc-18 homolog B) (Unc-18B) | -0.5779 | 3.62576 |
| CMPK2 | UMP-CMP kinase 2, mitochondrial (EC 2.7.4.14) (Nucleoside-diphosphate kinase) (EC 2.7.4.6) | 0.58505 | 2.30287 |
| CYBP | Calcyclin-binding protein (CacyBP) (hCacyBP) (S100A6-binding protein) (Siah-interacting protein) | 0.59727 | 2.10818 |
| NCF1B | Putative neutrophil cytosol factor 1B (NCF-1B) (Putative SH3 and PX domain-containing protein 1B) | 0.59912 | 2.59086 |
| NCF1 | Neutrophil cytosol factor 1 (NCF-1) (47 kDa autosomal chronic granulomatous disease protein) (47 kDa neutrophil oxidase factor) (NCF-47K) (Neutrophil NADPH oxidase factor 1) (Nox organizer 2) (Nox-organizing protein 2) (SH3 and PX domain-containing protein 1A) (p47-phox) | 0.59912 | 2.59086 |
| IGG1 | Immunoglobulin gamma-1 heavy chain (Immunoglobulin gamma-1 heavy chain NIE) | 0.6057 | 1.50324 |
| DDB2 | DNA damage-binding protein 2 (DDB p48 subunit) (DDBb) (Damage-specific DNA-binding protein 2) (UV-damaged DNA-binding protein 2) (UV-DDB 2) | 0.61114 | 2.99062 |
| PSB4 | Proteasome subunit beta type-4 (26 kDa prosomal protein) (HsBPROS26) (PROS-26) (Macropain beta chain) (Multicatalytic endopeptidase complex beta chain) (Proteasome beta chain) (Proteasome chain 3) (HsN3) | 0.61343 | 2.75272 |
| ATP5L | ATP synthase subunit g, mitochondrial (ATPase subunit g) (ATP synthase membrane subunit g) | 0.61531 | 1.56848 |
| ATX10 | Ataxin-10 (Brain protein E46 homolog) (Spinocerebellar ataxia type 10 protein) | 0.63205 | 2.66142 |
| PSB2 | Proteasome subunit beta type-2 (Macropain subunit C7-I) (Multicatalytic endopeptidase complex subunit C7-I) (Proteasome component C7-I) | 0.63278 | 2.23974 |
| GSTO1 | Glutathione S-transferase omega-1 (GSTO-1) (EC 2.5.1.18) (Glutathione S-transferase omega 1-1) (GSTO 1-1) (Glutathione-dependent dehydroascorbate reductase) (EC 1.8.5.1) (Monomethylarsonic acid reductase) (MMA(V) reductase) (EC 1.20.4.2) (S-(Phenacyl)glutathione reductase) (SPG-R) | 0.64442 | 1.72378 |
| SAM9L | Sterile alpha motif domain-containing protein 9-like (SAM domain-containing protein 9-like) | 0.64443 | 1.98439 |
| EST1 | Liver carboxylesterase 1 (Acyl-coenzyme A:cholesterol acyltransferase) (ACAT) (Brain carboxylesterase hBr1) (Carboxylesterase 1) (CE-1) (hCE-1) (EC 3.1.1.1) (Cholesteryl ester hydrolase) (CEH) (EC 3.1.1.13) (Cocaine carboxylesterase) (Egasyn) (HMSE) (Methylumbelliferyl-acetate deacetylase 1) (EC 3.1.1.56) (Monocyte/macrophage serine esterase) (Retinyl ester hydrolase) (REH) (Serine esterase 1) (Triacylglycerol hydrolase) (TGH) | 0.64642 | 3.02035 |
| DDX58 | DEAD box protein 58 (Antiviral innate immune response receptor RIG-1) | 0.64919 | 2.49513 |
| APEX1 | DNA-(apurinic or apyrimidinic site) endonuclease (EC 3.1.11.2) (APEX nuclease) (APEN) (Apurinic-apyrimidinic endonuclease 1) (AP endonuclease 1) (APE-1) (REF-1) (Redox factor-1) [Cleaved into: DNA-(apurinic or apyrimidinic site) endonuclease, mitochondrial] | 0.6559 | 1.31424 |
| A1AT | Alpha-1-antitrypsin (Alpha-1 protease inhibitor) (Alpha-1-antiproteinase) (Serpin A1) [Cleaved into: Short peptide from AAT (SPAAT)] | 0.65814 | 1.78611 |
| S10A8 | Protein S100-A8 (Calgranulin-A) (Calprotectin L1L subunit) (Cystic fibrosis antigen) (CFAG) (Leukocyte L1 complex light chain) (Migration inhibitory factor-related protein 8) (MRP-8) (p8) (S100 calcium-binding protein A8) (Urinary stone protein band A) | 0.67491 | 1.72197 |
| SAP | Prosaposin (Proactivator polypeptide) [Cleaved into: Saposin-A (Protein A); Saposin-B-Val; Saposin-B (Cerebroside sulfate activator) (CSAct) (Dispersin) (Sphingolipid activator protein 1) (SAP-1) (Sulfatide/GM1 activator); Saposin-C (A1 activator) (Co-beta-glucosidase) (Glucosylceramidase activator) (Sphingolipid activator protein 2) (SAP-2); Saposin-D (Component C) (Protein C)] | 0.67557 | 1.3805 |
| GCN1 | Stalled ribosome sensor GCN1 (GCN1 eIF-2-alpha kinase activator homolog) (GCN1-like protein 1) (General control of amino-acid synthesis 1-like protein 1) (Translational activator GCN1) (HsGCN1) | 0.68212 | 1.54991 |
| HNRL2 | Heterogeneous nuclear ribonucleoprotein U-like protein 2 (Scaffold-attachment factor A2) (SAF-A2) | 0.69468 | 1.44467 |
| DDX41 | Probable ATP-dependent RNA helicase DDX41 (EC 3.6.4.13) (DEAD box protein 41) (DEAD box protein abstrakt homolog) | 0.70937 | 2.3936 |
| KDM1A | Lysine-specific histone demethylase 1A (EC 1.14.99.66) (BRAF35-HDAC complex protein BHC110) (Flavin-containing amine oxidase domain-containing protein 2) ([histone H3]-dimethyl-L-lysine(4) FAD-dependent demethylase 1A) | 0.73111 | 1.61637 |
| TIAR | Nucleolysin TIAR (TIA-1-related protein) | 0.73732 | 2.06249 |
| CALR | Calreticulin (CRP55) (Calregulin) (Endoplasmic reticulum resident protein 60) (ERp60) (HACBP) (grp60) | 0.73835 | 3.33055 |
| MYL6 | Myosin light polypeptide 6 (17 kDa myosin light chain) (LC17) (Myosin light chain 3) (MLC-3) (Myosin light chain alkali 3) (Myosin light chain A3) (Smooth muscle and nonmuscle myosin light chain alkali 6) | 0.74997 | 1.75992 |
| CATD | Cathepsin D (EC 3.4.23.5) [Cleaved into: Cathepsin D light chain; Cathepsin D heavy chain] | 0.75227 | 2.02338 |
| TAP1 | Antigen peptide transporter 1 (APT1) (EC 7.4.2.14) (ATP-binding cassette sub-family B member 2) (Peptide supply factor 1) (Peptide transporter PSF1) (PSF-1) (Peptide transporter TAP1) (Peptide transporter involved in antigen processing 1) (Really interesting new gene 4 protein) (RING4) | 0.78453 | 2.82634 |
| CALX | Calnexin (IP90) (Major histocompatibility complex class I antigen-binding protein p88) (p90) | 0.79113 | 1.67873 |
| PSB10 | Proteasome subunit beta type-10 (EC 3.4.25.1) (Low molecular mass protein 10) (Macropain subunit MECl-1) (Multicatalytic endopeptidase complex subunit MECl-1) (Proteasome MECl-1) (Proteasome subunit beta-2i) | 0.80007 | 2.8627 |
| CATG | Cathepsin G (CG) (EC 3.4.21.20) [Cleaved into: Cathepsin G, C-terminal truncated form] | 0.80272 | 3.05875 |
| FILA | Filaggrin | 0.8126 | 1.58315 |
| PLS1 | Phospholipid scramblase 1 (PL scramblase 1) (Ca(2+)-dependent phospholipid scramblase 1) (Erythrocyte phospholipid scramblase) (Mg(2+)-dependent nuclease) (EC 3.1.-.-) (MmTRA1b) | 0.81489 | 3.94907 |
| ARK72 | Aflatoxin B1 aldehyde reductase member 2 (EC 1.1.1.n11) (AFB1 aldehyde reductase 1) (AFB1-AR 1) (Aldoketoreductase 7) (Succinic semialdehyde reductase) (SSA reductase) | 0.82628 | 1.36502 |
| TYPH | Thymidine phosphorylase (TP) (EC 2.4.2.4) (Gliostatin) (Platelet-derived endothelial cell growth factor) (PD-ECGF) (TdRPase) | 0.84238 | 2.5585 |
| THUM1 | THUMP domain-containing protein 1 | 0.85235 | 1.68893 |
| GRAH | Granzyme H (EC 3.4.21.-) (CCP-X) (Cathepsin G-like 2) (CTSGL2) (Cytotoxic T-lymphocyte proteinase) (Cytotoxic serine protease C) (CSP-C) | 0.90969 | 2.01037 |
| AMPL | Cytosol aminopeptidase (EC 3.4.11.1) (Cysteinylglycine-S-conjugate dipeptidase) (EC 3.4.13.23) (Leucine aminopeptidase 3) (LAP-3) (Leucyl aminopeptidase) (Peptidase S) (Proline aminopeptidase) (EC 3.4.11.5) (Prolyl aminopeptidase) | 0.9149 | 4.56312 |
| BST2 | Bone marrow stromal antigen 2 (BST-2) (HM1.24 antigen) (Tetherin) (CD antigen CD317) | 0.91809 | 2.00901 |
| TRFE | Serotransferrin (Transferrin) (Beta-1 metal-binding globulin) (Siderophilin) | 0.92589 | 1.30616 |
| SYWC | Tryptophan--tRNA ligase, cytoplasmic (EC 6.1.1.2) (Interferon-induced protein 53) (IFP53) (Tryptophanyl-tRNA synthetase) (TrpRS) (hWRS) [Cleaved into: T1-TrpRS; T2-TrpRS] | 0.95457 | 1.83803 |
| STAT1 | Signal transducer and activator of transcription 1-alpha/beta (Transcription factor ISGF-3 components p91/p84) | 0.9692 | 2.63354 |
| CD38 | ADP-ribosyl cyclase/cyclic ADP-ribose hydrolase 1 (EC 3.2.2.-) (EC 3.2.2.6) (2'-phospho-ADP-ribosyl cyclase) (2'-phospho-ADP-ribosyl cyclase/2'-phospho-cyclic-ADP-ribose transferase) (EC 2.4.99.20) (2'-phospho-cyclic-ADP-ribose transferase) (ADP-ribosyl cyclase 1) (ADPRC 1) (Cyclic ADP-ribose hydrolase 1) (cADPR hydrolase 1) (T10) (CD antigen CD38) | 0.9852 | 3.99935 |
| OAS2 | 2'-5'-oligoadenylate synthase 2 ((2-5')oligo(A) synthase 2) (2-5A synthase 2) (EC 2.7.7.84) (p69 OAS / p71 OAS) (p69OAS / p71OAS) | 1.00021 | 4.11193 |
| AACT | Alpha-1-antichymotrypsin (ACT) (Cell growth-inhibiting gene 24/25 protein) (Serpin A3) [Cleaved into: Alpha-1-antichymotrypsin His-Pro-less] | 1.02357 | 2.90383 |
| LG3BP | Galectin-3-binding protein (Basement membrane autoantigen p105) (Lectin galactoside-binding soluble 3-binding protein) (Mac-2-binding protein) (MAC2BP) (Mac-2 BP) (Tumor-associated antigen 90K) | 1.02765 | 3.55188 |
| IFIT2 | Interferon-induced protein with tetratricopeptide repeats 2 (IFIT-2) (ISG-54 K) (Interferon-induced 54 kDa protein) (IFI-54K) (P54) | 1.04792 | 3.96266 |
| 1A03 | HLA class I histocompatibility antigen, A alpha chain | 1.06703 | 1.37253 |
| ALBU | Albumin | 1.09259 | 1.32339 |
| TAP2 | Antigen peptide transporter 2 (APT2) (EC 7.4.2.14) (ATP-binding cassette sub-family B member 3) (Peptide supply factor 2) (Peptide transporter PSF2) (PSF-2) (Peptide transporter TAP2) (Peptide transporter involved in antigen processing 2) (Really interesting new gene 11 protein) (RING11) | 1.11687 | 3.3616 |
| OAS1 | 2'-5'-oligoadenylate synthase 1 ((2-5')oligo(A) synthase 1) (2-5A synthase 1) (EC 2.7.7.84) (E18/E16) (p46/p42 OAS) | 1.12718 | 2.7413 |
| TLN2 | Talin-2 | 1.15894 | 2.04804 |
| FBX6 | F-box only protein 6 (F-box protein that recognizes sugar chains 2) (F-box/G-domain protein 2) | 1.15963 | 3.0199 |
| PSB3 | Proteasome subunit beta type-3 (Proteasome chain 13) (Proteasome component C10-II) (Proteasome theta chain) | 1.27874 | 2.23182 |
| OAS3 | 2'-5'-oligoadenylate synthase 3 ((2-5')oligo(A) synthase 3) (2-5A synthase 3) (EC 2.7.7.84) (p100 OAS) (p100OAS) | 1.28759 | 4.33362 |
| IFIT1 | Interferon-induced protein with tetratricopeptide repeats 1 (IFIT-1) (Interferon-induced 56 kDa protein) (IFI-56K) (P56) | 1.35593 | 4.16924 |
| IFIT3 | Interferon-induced protein with tetratricopeptide repeats 3 (IFIT-3) (CIG49) (ISG-60) (Interferon-induced 60 kDa protein) (IFI-60K) (Interferon-induced protein with tetratricopeptide repeats 4) (IFIT-4) (Retinoic acid-induced gene G protein) (P60) (RIG-G) | 1.40951 | 4.86468 |
| MX2 | Interferon-induced GTP-binding protein Mx2 (Interferon-regulated resistance GTP-binding protein MxB) (Myxovirus resistance protein 2) (p78-related protein) | 1.4203 | 4.72113 |
| ISG15 | Ubiquitin-like protein ISG15 (Interferon-induced 15 kDa protein) (Interferon-induced 17 kDa protein) (IP17) (Ubiquitin cross-reactive protein) (hUCRP) | 1.5937 | 4.1169 |
| PARP9 | Protein mono-ADP-ribosyltransferase PARP9 (EC 2.4.2.-) (ADP-ribosyltransferase diphtheria toxin-like 9) (ARTD9) (B aggressive lymphoma protein) (Poly [ADP-ribose] polymerase 9) (PARP-9) | 1.65224 | 3.78095 |
| IFM2 | Interferon-induced transmembrane protein 2 (Dispanin subfamily A member 2c) (DSPA2c) (Interferon-inducible protein 1-8D) | 1.66974 | 3.31843 |
| IFM1 | Interferon-induced transmembrane protein 1 (Dispanin subfamily A member 2a) (DSPA2a) (Interferon-induced protein 17) (Interferon-inducible protein 9-27) (Leu-13 antigen) (CD antigen CD225) | 1.66974 | 3.31843 |
| IFM3 | Interferon-induced transmembrane protein 3 (Dispanin subfamily A member 2b) (DSPA2b) (Interferon-inducible protein 1-8U) | 1.66974 | 3.31843 |
| SN | Sialoadhesin (Sialic acid-binding Ig-like lectin 1) (Siglec-1) (CD antigen CD169) | 1.67598 | 3.94234 |
| TSP1 | Thrombospondin-1 (Glycoprotein G) | 1.77911 | 1.40843 |
| MX1 | Interferon-induced GTP-binding protein Mx1 (Interferon-induced protein p78) (IFI-78K) (Interferon-regulated resistance GTP-binding protein MxA) (Myxoma resistance protein 1) (Myxovirus resistance protein 1) [Cleaved into: Interferon-induced GTP-binding protein Mx1, N-terminally processed] | 2.00146 | 3.06241 |
| LBP | Lipopolysaccharide-binding protein (LBP) | 2.05851 | 1.84323 |
