## Supplementary table 2 for "Proteomics analysis of peripheral blood monocytes from patients in early dengue infection reveals potential markers of subsequent fluid leakage"

**Supplementary Table 2:** Differentially expressed proteins in DHF cell lysates compared to DF

| **Accession** | **Protein names** | **log_2_ fold change** | **-log_10_ p value** |
| --- | --- | --- | --- |
| FIBG | Fibrinogen gamma chain | -1.3631 | 1.47598 |
| NCKP1 | Nck-associated protein 1 (NAP 1) (Membrane-associated protein HEM-2) (p125Nap1) | -1.255 | 1.53176 |
| LMNB2 | Lamin-B2 | -1.1896 | 1.31612 |
| MMRN1 | Multimerin-1 (EMILIN-4) (Elastin microfibril interface located protein 4) (Elastin microfibril interfacer 4) (Endothelial cell multimerin) [Cleaved into: Platelet glycoprotein Ia*; 155 kDa platelet multimerin (p-155) (p155)] | -1.1584 | 3.05892 |
| VASP | Vasodilator-stimulated phosphoprotein (VASP) | -1.1429 | 1.44952 |
| ERAP2 | Endoplasmic reticulum aminopeptidase 2 (EC 3.4.11.-) (Leukocyte-derived arginine aminopeptidase) (L-RAP) | -1.1225 | 1.83566 |
| CAVN2 | Caveolae-associated protein 2 (Cavin-2) (PS-p68) (Phosphatidylserine-binding protein) (Serum deprivation-response protein) | -1.0455 | 1.64039 |
| URP2 | Fermitin family homolog 3 (Kindlin-3) (MIG2-like protein) (Unc-112-related protein 2) | -1.0264 | 3.12948 |
| SBDS | Ribosome maturation protein SBDS (Shwachman-Bodian-Diamond syndrome protein) | -1.0147 | 2.43506 |
| VWF | von Willebrand factor (vWF) [Cleaved into: von Willebrand antigen 2 (von Willebrand antigen II)] | -0.9961 | 1.53487 |
| ETFB | Electron transfer flavoprotein subunit beta (Beta-ETF) | -0.9822 | 1.4466 |
| RAB8A | Ras-related protein Rab-8A (EC 3.6.5.2) (Oncogene c-mel) | -0.9776 | 1.70114 |
| GTR14 | Solute carrier family 2, facilitated glucose transporter member 14 (Glucose transporter type 14) (GLUT-14) | -0.9664 | 3.17961 |
| GTR3 | Solute carrier family 2, facilitated glucose transporter member 3 (Glucose transporter type 3, brain) (GLUT-3) | -0.9664 | 3.17961 |
| ITB3 | Integrin beta-3 (Platelet membrane glycoprotein IIIa) (GPIIIa) (CD antigen CD61) | -0.9395 | 1.33539 |
| RAB1B | Ras-related protein Rab-1B (EC 3.6.5.2) | -0.9112 | 2.78043 |
| COPD | Coatomer subunit delta (Archain) (Delta-coat protein) (Delta-COP) | -0.8954 | 1.55949 |
| WASP | Actin nucleation-promoting factor WAS (Wiskott-Aldrich syndrome protein) (WASp) | -0.8855 | 2.51693 |
| CTGF | Cellular communication network family member 2 | -0.8791 | 1.43092 |
| TAGL2 | Transgelin-2 (Epididymis tissue protein Li 7e) (SM22-alpha homolog) | -0.8722 | 2.02525 |
| PSMD3 | 26S proteasome non-ATPase regulatory subunit 3 (26S proteasome regulatory subunit RPN3) (26S proteasome regulatory subunit S3) (Proteasome subunit p58) | -0.8666 | 1.31039 |
| MIC60 | MICOS complex subunit MIC60 (Cell proliferation-inducing gene 4/52 protein) (Mitochondrial inner membrane protein) (Mitofilin) (p87/89) | -0.855 | 3.11809 |
| TLN1 | Talin-1 | -0.8547 | 2.5701 |
| FIBA | Fibrinogen alpha chain [Cleaved into: Fibrinopeptide A; Fibrinogen alpha chain] | -0.8432 | 1.67125 |
| SNX2 | Sorting nexin-2 (Transformation-related gene 9 protein) (TRG-9) | -0.8399 | 3.43217 |
| DDX46 | Probable ATP-dependent RNA helicase DDX46 (EC 3.6.4.13) (DEAD box protein 46) (PRP5 homolog) | -0.8093 | 2.13341 |
| ILK | Integrin-linked protein kinase (EC 2.7.11.1) (59 kDa serine/threonine-protein kinase) (Beta-integrin-linked kinase) (ILK-1) (ILK-2) (p59ILK) | -0.8075 | 1.34749 |
| NHRF1 | Na(+)/H(+) exchange regulatory cofactor NHE-RF1 (NHERF-1) (Ezrin-radixin-moesin-binding phosphoprotein 50) (EBP50) (Regulatory cofactor of Na(+)/H(+) exchanger) (Sodium-hydrogen exchanger regulatory factor 1) (Solute carrier family 9 isoform A3 regulatory factor 1) | -0.8033 | 2.19932 |
| SYYC | Tyrosine--tRNA ligase, cytoplasmic (EC 6.1.1.1) (Tyrosyl-tRNA synthetase) (TyrRS) [Cleaved into: Tyrosine--tRNA ligase, cytoplasmic, N-terminally processed] | -0.8001 | 1.64877 |
| GNAQ | Guanine nucleotide-binding protein G(q) subunit alpha (Guanine nucleotide-binding protein alpha-q) | -0.7853 | 1.60054 |
| OPA1 | Dynamin-like 120 kDa protein, mitochondrial (EC 3.6.5.5) (Optic atrophy protein 1) [Cleaved into: Dynamin-like 120 kDa protein, form S1] | -0.7764 | 2.01221 |
| PARVB | Beta-parvin (Affixin) | -0.7763 | 1.51957 |
| GPV | Platelet glycoprotein V (GPV) (Glycoprotein 5) (CD antigen CD42d) | -0.7716 | 3.61414 |
| SEPT6 | Septin-6 | -0.7716 | 1.58305 |
| NSF1C | NSFL1 cofactor p47 (UBX domain-containing protein 2C) (p97 cofactor p47) | -0.7666 | 1.75665 |
| GP1BA | Platelet glycoprotein Ib alpha chain (GP-Ib alpha) (GPIb-alpha) (GPIbA) (Glycoprotein Ibalpha) (Antigen CD42b-alpha) (CD antigen CD42b) [Cleaved into: Glycocalicin] | -0.7512 | 1.31745 |
| ANXA7 | Annexin A7 (Annexin VII) (Annexin-7) (Synexin) | -0.7509 | 3.05138 |
| EZRI | Ezrin (Cytovillin) (Villin-2) (p81) | -0.7503 | 1.70886 |
| RDH11 | Retinol dehydrogenase 11 (EC 1.1.1.300) (Androgen-regulated short-chain dehydrogenase/reductase 1) (HCV core-binding protein HCBP12) (Prostate short-chain dehydrogenase/reductase 1) (Retinal reductase 1) (RalR1) (Short chain dehydrogenase/reductase family 7C member 1) | -0.7502 | 3.18262 |
| RHOF | Rho-related GTP-binding protein RhoF (Rho family GTPase Rif) (Rho in filopodia) | -0.7496 | 1.55845 |
| SF3A3 | Splicing factor 3A subunit 3 (SF3a60) (Spliceosome-associated protein 61) (SAP 61) | -0.7466 | 2.1756 |
| AT2A3 | Sarcoplasmic/endoplasmic reticulum calcium ATPase 3 (SERCA3) (SR Ca(2+)-ATPase 3) (EC 7.2.2.10) (Calcium pump 3) | -0.7256 | 2.60096 |
| ICAM2 | Intercellular adhesion molecule 2 (ICAM-2) (CD antigen CD102) | -0.7204 | 1.78744 |
| RALB | Ras-related protein Ral-B (EC 3.6.5.2) | -0.7122 | 2.17143 |
| GCN1 | Stalled ribosome sensor GCN1 (GCN1 eIF-2-alpha kinase activator homolog) (GCN1-like protein 1) (General control of amino-acid synthesis 1-like protein 1) (Translational activator GCN1) (HsGCN1) | -0.7098 | 1.35645 |
| RB27B | Ras-related protein Rab-27B (EC 3.6.5.2) (C25KG) | -0.7075 | 1.44913 |
| IF4A2 | Eukaryotic initiation factor 4A-II (eIF-4A-II) (eIF4A-II) (EC 3.6.4.13) (ATP-dependent RNA helicase eIF4A-2) | -0.7053 | 1.3219 |
| CIP4 | Cdc42-interacting protein 4 (Protein Felic) (Salt tolerant protein) (hSTP) (Thyroid receptor-interacting protein 10) (TR-interacting protein 10) (TRIP-10) | -0.7047 | 1.63399 |
| ITA2B | Integrin alpha-IIb (GPalpha IIb) (GPIIb) (Platelet membrane glycoprotein IIb) (CD antigen CD41) [Cleaved into: Integrin alpha-IIb heavy chain; Integrin alpha-IIb light chain, form 1; Integrin alpha-IIb light chain, form 2] | -0.7035 | 2.05273 |
| ECE1 | Endothelin-converting enzyme 1 (ECE-1) (EC 3.4.24.71) | -0.6973 | 2.03304 |
| EHD3 | EH domain-containing protein 3 (PAST homolog 3) | -0.6893 | 1.68911 |
| F13A | Coagulation factor XIII A chain (Coagulation factor XIIIa) (EC 2.3.2.13) (Protein-glutamine gamma-glutamyltransferase A chain) (Transglutaminase A chain) | -0.6787 | 2.85746 |
| HSPB1 | Heat shock protein beta-1 (HspB1) (28 kDa heat shock protein) (Estrogen-regulated 24 kDa protein) (Heat shock 27 kDa protein) (HSP 27) (Stress-responsive protein 27) (SRP27) | -0.6697 | 1.38016 |
| RINI | Ribonuclease inhibitor (Placental ribonuclease inhibitor) (Placental RNase inhibitor) (Ribonuclease/angiogenin inhibitor 1) (RAI) | -0.6695 | 1.49174 |
| COR1C | Coronin-1C (Coronin-3) (hCRNN4) | -0.6543 | 1.33242 |
| AT2A2 | Sarcoplasmic/endoplasmic reticulum calcium ATPase 2 (SERCA2) (SR Ca(2+)-ATPase 2) (EC 7.2.2.10) (Calcium pump 2) (Calcium-transporting ATPase sarcoplasmic reticulum type, slow twitch skeletal muscle isoform) (Endoplasmic reticulum class 1/2 Ca(2+) ATPase) | -0.6473 | 2.90478 |
| ITB1 | Integrin beta-1 (Fibronectin receptor subunit beta) (Glycoprotein IIa) (GPIIA) (VLA-4 subunit beta) (CD antigen CD29) | -0.6395 | 1.9509 |
| ROA2 | Heterogeneous nuclear ribonucleoproteins A2/B1 (hnRNP A2/B1) | -0.6334 | 1.61068 |
| TPM4 | Tropomyosin alpha-4 chain (TM30p1) (Tropomyosin-4) | -0.6312 | 1.32585 |
| RL28 | Large ribosomal subunit protein eL28 (60S ribosomal protein L28) | -0.6254 | 1.33041 |
| BAP31 | B-cell receptor-associated protein 31 (BCR-associated protein 31) (Bap31) (6C6-AG tumor-associated antigen) (Protein CDM) (p28) | -0.6241 | 2.68604 |
| COPA | Coatomer subunit alpha (Alpha-coat protein) (Alpha-COP) (HEP-COP) (HEPCOP) [Cleaved into: Xenin (Xenopsin-related peptide); Proxenin] | -0.6241 | 2.44365 |
| EM55 | 55 kDa erythrocyte membrane protein (p55) (Membrane protein, palmitoylated 1) | -0.6157 | 1.38376 |
| HS90B | Heat shock protein HSP 90-beta (HSP 90) (Heat shock 84 kDa) (HSP 84) (HSP84) | -0.6156 | 1.32305 |
| IRAK4 | Interleukin-1 receptor-associated kinase 4 (IRAK-4) (EC 2.7.11.1) (Renal carcinoma antigen NY-REN-64) | -0.613 | 1.34218 |
| PAK2 | Serine/threonine-protein kinase PAK 2 (EC 2.7.11.1) (Gamma-PAK) (PAK65) (S6/H4 kinase) (p21-activated kinase 2) (PAK-2) (p58) [Cleaved into: PAK-2p27 (p27); PAK-2p34 (p34) (C-t-PAK2)] | -0.6001 | 2.00303 |
| ATP5L | ATP synthase subunit g, mitochondrial (ATPase subunit g) (ATP synthase membrane subunit g) | -0.5998 | 1.76212 |
| G6PD | Glucose-6-phosphate 1-dehydrogenase (G6PD) (EC 1.1.1.49) | -0.5983 | 2.29229 |
| RS3A | Small ribosomal subunit protein eS1 (40S ribosomal protein S3a) (v-fos transformation effector protein) (Fte-1) | -0.597 | 1.46349 |
| KAPCA | cAMP-dependent protein kinase catalytic subunit alpha (PKA C-alpha) (EC 2.7.11.11) | -0.5939 | 2.01732 |
| NID1 | Nidogen-1 (NID-1) (Entactin) | -0.5915 | 2.44218 |
| CLH1 | Clathrin heavy chain 1 (Clathrin heavy chain on chromosome 17) (CLH-17) | -0.5899 | 2.45898 |
| DEK | Protein DEK | -0.5893 | 1.97392 |
| ATP5I | ATP synthase subunit e, mitochondrial (ATPase subunit e) (ATP synthase membrane subunit e) [Cleaved into: ATP synthase subunit e, mitochondrial, N-terminally processed] | -0.5873 | 1.36113 |
| RSU1 | Ras suppressor protein 1 (RSP-1) (Rsu-1) | -0.5843 | 1.7062 |
| TCPZ | T-complex protein 1 subunit zeta (TCP-1-zeta) (Acute morphine dependence-related protein 2) (CCT-zeta-1) (HTR3) (Tcp20) | -0.5811 | 1.34918 |
| FLNA | Filamin-A (FLN-A) (Actin-binding protein 280) (ABP-280) (Alpha-filamin) (Endothelial actin-binding protein) (Filamin-1) (Non-muscle filamin) | -0.5795 | 2.14458 |
| MK01 | Mitogen-activated protein kinase 1 (MAP kinase 1) (MAPK 1) (EC 2.7.11.24) (ERT1) (Extracellular signal-regulated kinase 2) (ERK-2) (MAP kinase isoform p42) (p42-MAPK) (Mitogen-activated protein kinase 2) (MAP kinase 2) (MAPK 2) | -0.5775 | 2.01257 |
| K1C9 | Keratin, type I cytoskeletal 9 (Cytokeratin-9) (CK-9) (Keratin-9) (K9) | 0.59726 | 1.99881 |
| HORN | Hornerin | 0.67201 | 2.98874 |
| PSB2 | Proteasome subunit beta type-2 (Macropain subunit C7-I) (Multicatalytic endopeptidase complex subunit C7-I) (Proteasome component C7-I) | 0.70867 | 1.53764 |
| K22E | Keratin, type II cytoskeletal 2 epidermal (Cytokeratin-2e) (CK-2e) (Epithelial keratin-2e) (Keratin-2 epidermis) (Keratin-2e) (K2e) (Type-II keratin Kb2) | 0.74177 | 1.43604 |
| NGAL | Neutrophil gelatinase-associated lipocalin (NGAL) (25 kDa alpha-2-microglobulin-related subunit of MMP-9) (Lipocalin-2) (Oncogene 24p3) (Siderocalin) (p25) | 0.76008 | 2.44488 |
| PSB3 | Proteasome subunit beta type-3 (Proteasome chain 13) (Proteasome component C10-II) (Proteasome theta chain) | 0.82007 | 1.31385 |
| NCF1 | Neutrophil cytosol factor 1 (NCF-1) (47 kDa autosomal chronic granulomatous disease protein) (47 kDa neutrophil oxidase factor) (NCF-47K) (Neutrophil NADPH oxidase factor 1) (Nox organizer 2) (Nox-organizing protein 2) (SH3 and PX domain-containing protein 1A) (p47-phox) | 0.82749 | 2.50284 |
| NCF1B | Putative neutrophil cytosol factor 1B (NCF-1B) (Putative SH3 and PX domain-containing protein 1B) | 0.82749 | 2.50284 |
| ARI2 | E3 ubiquitin-protein ligase ARIH2 (ARI-2) (Protein ariadne-2 homolog) (EC 2.3.2.31) (RING-type E3 ubiquitin transferase ARIH2) (Triad1 protein) | 0.82933 | 1.72245 |
| S10A9 | Protein S100-A9 (Calgranulin-B) (Calprotectin L1H subunit) (Leukocyte L1 complex heavy chain) (Migration inhibitory factor-related protein 14) (MRP-14) (p14) (S100 calcium-binding protein A9) | 0.85903 | 1.93371 |
| S10A8 | Protein S100-A8 (Calgranulin-A) (Calprotectin L1L subunit) (Cystic fibrosis antigen) (CFAG) (Leukocyte L1 complex light chain) (Migration inhibitory factor-related protein 8) (MRP-8) (p8) (S100 calcium-binding protein A8) (Urinary stone protein band A) | 0.93615 | 1.76402 |
| BPI | Bactericidal permeability-increasing protein (BPI) (CAP 57) | 0.98905 | 1.69293 |
